# Blood Pressure Reduction in Multiomic Digital Food-as-Medicine: a single-arm cohort study with a regression-to-the-mean-adjusted estimand

**DOI:** 10.64898/2026.09.24.26363892

**Authors:** Inti Pedroso, Shreyas V. Kumbhare, Santosh K. Saravanan, Garima Sharma, Daniel E. Almonacid, Ranjan Sinha

**Affiliations:** Digbi Health, CA, USA

**Keywords:** home blood pressure monitoring, hypertension, regression to the mean, digital health, real-world evidence, food as medicine, negative control population, single-arm cohort

## Abstract

Elevated systolic blood pressure (SBP) is a major modifiable driver of cardiovascular morbidity. Digital food-as-medicine and home-blood-pressure-monitoring programs increasingly support its management outside the clinic, but most published evaluations are single-arm, pre/post designs; because enrolment is conditioned on an elevated baseline reading, regression to the mean (RTM) is expected to inflate any observed improvement, and this is rarely checked against an internal comparator. We conducted a retrospective, single-arm cohort study of home blood-pressure monitoring within a commercial digital food-as-medicine program serving a US employer-sponsored population with obesity. Of 329 members included, 136 were at-goal (<130/80 mmHg), 74 at Stage 1 (130–134/80–84 mmHg) and 119 at Stage 2 (≥135/85 mmHg) at baseline; follow-up SBP was evaluable for 128, 68 and 112 members in these strata at 3 months and for 62, 34 and 53 at 6 months, respectively. The primary estimand was the unadjusted within-member change in home SBP at 6 months among members with Stage-1 and Stage-2 blood pressure at baseline; a secondary estimand corrected for RTM using an ANCOVA linear mixed model with the at-goal stratum as an internal negative-control population. Among Stage-1 members (n = 34), unadjusted SBP change was −4.36 mmHg (95% CI −7.43 to −1.28; p = 0.0069), paired with an RTM-corrected secondary change of −3.33 mmHg (−6.40 to −0.26; p = 0.033). Among Stage-2 members (n = 53), unadjusted change was −10.17 mmHg (−12.94 to −7.40; p < 0.001), paired with an RTM-corrected secondary change of −5.50 mmHg (95% CI −8.50 to −2.50; p = 0.00037); RTM accounted for ∼24% and ∼46% of the unadjusted decline in Stage-1 and Stage-2, respectively. RTM-corrected estimates were robust to sensitivity analyses on measurement modality, seasonality, covariates and model specification, with no evidence of attrition bias. Program participation was associated with a decline in home SBP among Stage-1 and Stage-2 members that remained significant, though smaller, after RTM correction, supporting RTM-adjusted SBP change as the appropriate metric for evaluating food-as-medicine interventions.

## 2 Introduction

Elevated systolic blood pressure (SBP) remains among the largest modifiable contributors to cardiovascular morbidity, and the benefit attached to lowering it is well characterised. In a meta-analysis of randomised trials, every 10 mmHg reduction in systolic blood pressure was associated with a relative risk of 0.80 (95% CI 0.77–0.83) for major cardiovascular events, 0.73 (0.68–0.77) for stroke, 0.83 (0.78–0.88) for coronary heart disease and 0.72 (0.67–0.78) for heart failure (1). In a second meta-analysis of pharmacological interventions reported that a 5 mmHg decrease in SBP was associated with a hazard ratio for major cardiovascular events of 0.91 (95% CI 0.89–0.94) in participants without prior cardiovascular disease and 0.89 (0.86–0.92) in those with it (2). This disease burden reduction associated with decrease in SBP sets an informative background to which compare potential impact of BP interventions.

Home blood pressure monitoring is recommended by hypertension-society guidelines as the preferred modality for assessing control between clinic visits (3), and digital, coaching-based blood pressure programs have become a common vehicle for delivering hypertension management at scale, alongside food-as-medicine interventions that address the same outcome through diet. Published evaluations of digital blood-pressure programs and food-as-medicine interventions delivered without a concurrent control are predominantly single-arm, within-person pre/post comparisons, and uncorrected reductions in the range of 7 to 10 mmHg in home-monitored more severely elevated subgroups. For instance, a home-monitoring pilot reported −10.3 mmHg (95% CI –13.4 to –7.1) for SBP among members with baseline systolic pressure ≥130/≥80 mmHg (4), and a follow-up study in a larger sample reported −8.1 mmHg (95% CI –9.0 to –7.1) for the same patient group (5). An AI-guided digital intervention reported a change of –14.2 mmHg (95% CI −17.7 to −10.7) (6), and a multisite produce-prescription program reported −8.38 mmHg (95% CI −10.13 to −6.62; (7)). In contrast randomized studies with a pooled comparator-arm evidence, in which a concurrent or between-arm design cancels shared regression to the mean, reports a wider and generally smaller range of net effects: −11.4 mmHg (95% CI −15.9 to −6.9) for a fully controlled-feeding combination diet against a control diet among participants with hypertension (8), a net 4.3 mmHg (95% CI −5.9 to −2.8) for a behavioural plus Dietary Approach to Stop Hypertension (DASH) arm against advice-only at 6 months (9), a −5.5 mmHg (95% CI 1.3–9.8) SBP between-arm difference at 13 weeks in which the control arm, which received only a BP monitor and no coaching, itself fell 7.1 mmHg (10), and a pooled effect of −2.76 mmHg (95% CI −3.94 to −1.58) at 6 months across 76 comparator-arm studies (11).

The difference in magnitude of SBP change reported by randomized trials and single arm is largely associated with the RTM. Entry into a blood pressure program is conditioned on an elevated reading, so the baseline value is selected upward and any subsequent measurement regresses toward the individual’s underlying mean irrespective of the intervention. RTM is a well-documented challenge for evidence of blood pressure change collected before and after an intervention with no concurrent control group (12–15), and methods that estimate and remove the baseline regression component of an observed change without a concurrent control arm are established (13,16–18). Beyond the reach and scale that a purely digital delivery channel offers, evidence on the cost-effectiveness of digital health interventions for chronic disease management is favourable with the majority of rigorously evaluated programs found cost-effective or cost-saving (19,20).

The digital health program evaluated in this study is not built around a hypertension-specific, guideline-referenced dietary protocol, and does not prescribe DASH or a sodium target as such. It delivers a food-as-medicine curriculum whose individual components nonetheless overlap substantially with the dietary patterns tested in the hypertension literature: reduced refined carbohydrate and ultra-processed food intake, reduced alcohol, increased dietary fibre and vegetable diversity, and structured weight loss delivered through a CDC-recognised Diabetes Prevention Program (DPP+) curriculum. Each of these levers carries an independent randomised or meta-analytic evidence base for blood pressure lowering, with effects consistently concentrated in participants who are already hypertensive. Weight loss shows a dose-response of −1.05 mmHg SBP per kilogram lost (95% CI −1.43 to −0.66) across 25 randomised trials, and roughly twice the reduction above a 5 kg loss threshold (21,22). Real-world translations of the DPP model pooled −4.29 mmHg SBP (95% CI −5.73 to −2.84) across 44 community programs, and −4.62 mmHg (95% CI −6.48 to −2.76) in the subset with a maintenance phase (23). While Orchard et al. (24) showed a significant reduction of high BP among at three years follow up for participants in the DPP lifestyle intervention with or without metabolic syndrome at baseline. Reduced alcohol intake lowers SBP by −3.31 mmHg with a clear dose-response replicated across non-overlapping meta-analyses of 15 and 36 trials (25,26). Increased fibre and vegetable intake produces only modest overall change (−1.15 mmHg SBP, non-significant) but −5.95 mmHg (95% CI −9.50 to −2.40) in hypertensive participants, and − 2.5 mmHg to −4.3 mmHg in trials targeting hypertension management (8,27–30). Higher ultra-processed food intake is associated with incident hypertension in large prospective cohorts (31,32), but randomized trial evidence has yet to show that reducing ultra-processed food intake lowers blood pressure.

This program has also been evaluated directly on outcomes upstream of blood pressure. Previous studies have documented clinically meaningful impact of the digital health intervention for weight loss, improved glycemic control, reduced gastrointestinal symptoms, reduced mental health symptoms and improved microbiome profiles in real-world employer populations (33–38). Given the weight reductions achieved in those cohorts and the per-kilogram dose-response above, a blood pressure decline of a few mmHg is a plausible expectation for this program, with a larger decline plausible in participants entering with elevated readings. That expectation, rather than a hypertension-specific protocol, is what the present analysis sets out to test, including regression to the mean correction.

In this study we describe a single-arm, within-subject home blood pressure change across baseline strata in a contemporary cohort of a digital food-as-medicine program. Our objective is to estimate the unadjusted change alongside a RTM-corrected estimate.

## 3 Methodology

### 3.1 Study design and setting

This was a retrospective, single-arm study using self-reported, routinely collected application and connected-device data from a commercial digital food-as-medicine program operating in the United States.

The study’s primary outcome was change in home SBP from the baseline window to the 6-month landmark. Secondary outcomes were change in home SBP at the 3-month landmark and change in home diastolic pressure at both landmarks. Outcomes were analysed for the full cohort and stratified by baseline SBP stratum, namely at-goal, Stage 1 and Stage 2 which are defined later in this article. Change in within-person body weight over the same interval was analysed as a correlate of home SBP.

Completed STROBE for cohort studies (39) with the RECORD extension for studies using routinely collected health data (40) are provided as Supplementary Material.

### 3.2 Participants and digital health intervention

Program members, employees and dependents, enrolled into the digital health platform through their employer-sponsored benefits if they complied with the inclusion criteria: adults with ≥18 years of age (mandatory inclusion criterion), residing in the United States at the time of the study, with BMI > 30 or > 27 with an obesity related comorbidity, and had access to a compatible mobile device with internet connection. For this study we focus on the subset of members who self-reported a hypertension medical diagnosis and provided valid baseline home blood pressure readings (see below). The inclusion criteria further restricts to members that enrolled from October 2025 onwards, through 31 August 2026, which comprises a homogenous subset in terms of blood-pressure measurement device (see below) and contemporary effects, as compared to the platform historical cohort which is mostly from connected wearable devices.

The Institutional Review Board of E&I Review Services reviewed and approved the study protocol (protocol code #18053, approved 05/22/2018). Before being included in the study, all participants provided electronic informed consent as part of their written consent process.

Eligibility was assessed against a source population of members with at least one home blood pressure reading recorded in the platform’s data feed, after de-duplication and a plausibility filter. Each member’s index date (t0) was defined as the calendar day of their app download. Four analytic exclusion rules were then applied hierarchically, in a fixed order, so that each member is counted under only the first rule that excluded them: insufficient baseline monitoring (fewer than three days of baseline readings); no evaluable follow-up landmark by the observation cutoff, i.e. administratively censored before enough calendar time had elapsed; an evaluable follow-up window that was open but contained no observed reading; and an index date falling outside the cohort’s enrolment window.

The program is a digital therapeutics platform that combines baseline participant information collected through a health questionnaire with continuous monitoring delivered through an AI-powered mobile application: participants received a cellular– or Bluetooth-connected weight scale and, depending of the payer’s coverage, those self-reporting hypertension receive a cellular– or Bluetooth-connected upper-arm blood pressure cuff, and used the application to receive educational material and content and to log dietary intake via uploaded food photographs, which an AI-based meal rating and recommendation system scored across three dietary-harm axes, insufficient fiber, inflammatory potential, and expected insulin response, to inform recommendations to improve their meals (see details in Supplementary Methods).

### 3.3 Data collection and analysis

Members were provided with a Tenovi Cellular Blood Pressure Monitor (www.tenovi.com) together with instructions for its set up and use. Members connected the device to the mobile phone and to the program’s mobile app. In absence of a cellular blood pressure monitor, e.g., not compatible with the employer’s benefits, members provided blood pressure information either manually or through wearable device connectivity (Apple Health Kit or Google Fit). Readings arriving through a phone health-data integration (Apple HealthKit or Google Fit) or manual in-app entry come from member-owned devices; the robustness of the results to measurement modality was assessed in dedicated sensitivity analyses (see Sensitivity analyses).

A physiological plausibility filter removed values outside SBP 70–260 mmHg or DBP 40–160 mmHg before aggregation, a device– or entry-error guard rather than a clinical threshold; no distribution based trimming was applied, consistent with home-monitoring guidance that extreme-but-plausible readings can carry real signal (3,41).

Blood pressure readings were then aggregated in three stages so that members are not weighted by measurement frequency. Individual readings within a member were first clustered into “occasions”, a maximal run of readings no more than 15 minutes apart which approximates clinical practice of taking two to three readings in succession per sitting. These “occasions” were averaged to ‘per-day means’, and ‘per-day means’ were averaged to ‘window means’.

The baseline window spanned the 30 days from each member’s app-download date, following the convention of averaging measurements closest to program start and up to 30 days after (5). Baseline sufficiency required at least three monitoring occasions spread across at least three distinct days, or at least four occasions across at least two distinct days. Landmark windows were defined at 3 months (90 days ±30 days, formally from 60 to 120 days) and 6 months (180 days ±30 days, formally from 150 to 210 days). Unlike the baseline window, no minimum-days requirement was applied to a landmark window: a single observed reading falling within a landmark window was sufficient for that member to contribute an evaluable observation at that landmark.

Members were stratified based on home monitoring thresholds rather than office thresholds: at-goal, home <130/80 mmHg (office equivalent <130/80); Stage 1, home 130–134/80–84 mmHg (office equivalent approximately 130–139/80–89); and Stage 2, home ≥135/85 mmHg (office equivalent approximately ≥140/90) (3,41,42). As per the guidelines stratum assignment used an ‘*OR*’ rule on the two baseline readings, evaluated in a fixed order: Stage 2 required either baseline SBP ≥135 mmHg or DBP ≥85 mmHg; Stage 1 required either SBP ≥130 mmHg or DBP ≥80 mmHg; membership was at-goal otherwise. No rounding was applied to non-integer baseline means; classification used each member’s raw baseline value directly.

We used self-administered questionnaires to collect baseline information on sex, age, disease comorbidities including Type 2 Diabetes (T2D) and Chronic Kidney Disease (CKD), and self-reported blood markers including A1C, fasting glucose, LDL, HDL, total triglycerides, and hsCRP, and self-reported medication use.

### 3.4 Statistical analysis

#### 3.4.1 Models and estimands

The study’s primary estimand is the unadjusted mean paired change (within-member change) in home blood pressure from the baseline window to the landmark window, for members in the Stage-1 and Stage-2 stratum at month 6 with confidence interval with a t-distribution. The study’s secondary estimand is the RTM-corrected mean change in home blood pressure from baseline within a given baseline stratum at a given landmark, the model-estimated marginal mean (least-squares mean) for that stratum × landmark cell, evaluated at the cohort grand-mean baseline blood pressure. It is estimated by a reduced ANCOVA linear mixed model: change in home blood pressure regressed on centred baseline blood pressure, with a stratum × landmark interaction and a member-level random intercept, and nothing else: 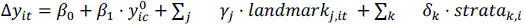 + 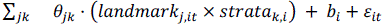, where Δ*y_it_* is the change in outcome *y* (SBP or DBP) for member *i* at landmark *t* relative to that member’s own baseline window; 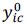 is member *i*’s baseline value of the same outcome, centred on the cohort mean; *landmark*_j,*it*_ and *strata_k_*_,*i*_ are indicator (dummy) codings of landmark (month 3 as reference strata) and home-BP stratum (at goal (reference strata), Stage 1 and Stage 2); *b_i_*∼ *N*(0, *τ*^2^) is the member-level random intercept; and *ε_it_*∼ *N*(0, *σ*^2^) is the residual. The model is fitted by restricted maximum likelihood using R (43) with lme4 (44) and emmeans (45). *β*_1_ is the coefficient on the centred baseline, reported as the regression-to-the-mean slope.

The at-goal stratum was designated in advance of estimation as an, and hypothesized to be behave like a valid, internal negative control population under the logic that members already at goal have limited room to improve, so if regression to the mean is operating, this stratum should show an apparent rise before adjustment, as its below-mean baseline readings regress upward toward the population mean, and that apparent rise should be attenuated by the same RTM correction (17,18).

#### 3.4.2 Sensitivity analyses

We performed sensitivity analyses with a covariate-adjusted model, the RTM-adjusted model including age (grand-mean-centred), baseline BMI category, self-reported type 2 diabetes, chronic kidney disease and baseline antihypertensive status. Estimated marginal means (EMMs) were formed as design-row linear combinations evaluated at each cell’s own covariate composition, at centred baseline SBP = 0. The RTM model requires no covariate imputation and was fitted on all eligible member × landmark rows, whereas the covariate-adjusted model was fitted on members with every covariate observed, so that the drop-one comparisons below vary the specification against a fixed sample. BMI categories were defined as per World Health Organization classification: normal weight (18.5–24.9 kg/m²), overweight (25.0–29.9 kg/m²), Class I obesity (30.0–34.9 kg/m²), Class II obesity (35.0–39.9 kg/m²), and Class III obesity (≥ 40.0 kg/m²). The covariate-adjusted model was refitted omitting age, self-reported T2D, CKD and baseline antihypertensive status in turn (BMI category retained in each), and omitting all five together, on a fixed complete-case sample, so that each drop-one row differs from the others only in specification and not in who is analysed.

The proportion of the unadjusted change attributable to regression to the mean was estimated as 1 − (RTM-model EMM ÷ unadjusted mean change), with confidence intervals by the multivariate delta method.

To evaluate the sensitivity of the main results to the blood pressure measurement modality we carried out three analyses: the RTM model refitted on device-provided members alone; the same model refitted on all non-device-provided members; and the RTM model on the full sample with modality entered as a fixed effect. Members were classified as “device-provided” (a cellular upper-arm cuff was dispatched to the member), “phone-integrated” (readings arriving only through a phone-side health-data integration, no dispatched cuff), “manual entry” (in-app keyed entry only), or “mixed”. A member with any dispatched cuff is device-provided for all of that member’s readings, including any keyed before dispatch.

Three further sensitivities extended the reduced RTM model itself. A season/calendar-month sensitivity added *sine* and *cosine* harmonic terms for calendar month to the reduced model’s formula, to test whether secular or seasonal blood-pressure variation confounds the RTM-corrected estimate. A stratum × baseline interaction sensitivity added a centred-baseline × stratum interaction term to the reduced model and tested it by a likelihood-ratio test comparing nested maximum likelihood fits, evaluating whether the pooled regression-to-the-mean slope differs across strata (16,46). An alternative RTM correction was formed from the at-goal (control) stratum’s own baseline-to-change slope, fitted on that stratum alone and applied to the other strata’s unadjusted cell means by the delta method (covariance assumed zero), as an alternative to the pooled-slope correction.

We assessed robustness to missing follow-up and informative attrition using three complementary estimators of mean SBP change from baseline: a delta-adjusted pattern-mixture model (47), inverse-probability-of-censoring weighting (IPCW; (48)), and an unweighted naive complete-case estimate as a reference point. All three were fitted at 3 and 6 month landmarks, separately within each home-BP stratum (Stage 1, Stage 2). Each estimates change by ANCOVA on centred baseline SBP; the IPCW censoring model used age, BMI category, type 2 diabetes, chronic kidney disease, baseline antihypertensive status and device provision. The pattern-mixture estimator treats each member’s unobserved follow-up SBP change as missing rather than dropping the member, and imputes it by conditional-mean imputation from a mixed model for repeated measures of the 3– and 6-month changes conditional on centred baseline SBP (rbmi R package; jackknife standard errors), estimating each stratum’s mean change by ANCOVA on centred baseline SBP; the base-case (δ = 0) estimate assumes these imputed values are missing at random. A tipping-point analysis (51) then re-fit the pooled estimate after adding a fixed erosion delta to every imputed (not observed) value only, swept over a grid from 0 to 8 mmHg in the direction that erodes the observed decline toward the null; the tipping δ reported for each stratum/landmark cell is the smallest delta in this grid at which the 95% CI first includes zero, so δ = 0 means the estimate is already non-significant before any erosion is applied. IPCW instead reweights members actually observed at a landmark by the inverse of their estimated probability of remaining uncensored, modelled from these covariates, with missing covariates multiply imputed by chained equations (49) and pooled by Rubin’s rules (50); the naive complete-case estimate applies neither imputation nor reweighting, restricting to members observed at that landmark. Sample size accordingly differs by estimator within a cell: the pattern-mixture n is the fixed size of the imputation panel for that stratum (constant across landmarks, since members are imputed rather than dropped), the IPCW ‘n’ is a Kish-effective weighted sample size, and the naive n is the literal count of members observed at that landmark.

Independently of the outcome regression, an RTM expectation for the change score was computed from within-member measurement reliability alone: ρ, the intraclass correlation of the baseline home systolic reading, was estimated as intra-class corrrelation ICC(1) from a random-intercept model fitted by restricted miximum-likelihood (REML) on day-level means within the same baseline window (0–30 days) that anchors the RTM features, using this cohort’s own reading-level home-monitoring data for members with a day-level baseline-window reading, an earlier-funnel-stage larger set than the primary analytic sample with sufficient baseline readings, drawn on for precision in this nuisance parameter alone. The classical regression-to-the-mean expectation for the change score is then E[X₂ − X₁] = −(1 − ρ)(xˉ_stratum − μ), evaluated at each stratum’s own mean centred baseline SBP (one row per member). The negative of the textbook level-prediction formula, because subtracting the baseline flips its sign (52). The reliability-based figure is a point estimate only, with no sampling variability propagated through it, while its model-implied comparator carries a 95% CI (normal reference distribution).

As an exploratory analysis, we tested whether members’ weight change and systolic change shared a common within– and between-member relationship, by decomposing the weight-change predictor into a within-member term (a member’s own bi-weekly weight change, expressed as a deviation from that member’s average) and a between-member term (that member’s average weight change, expressed as a deviation from the cohort average) using a Mundlak for predictor *x_it_* (member (i), bi-weekly period 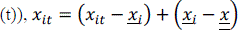, where 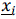 is member (i)’s own mean and 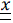 is the mean of person means (not the row grand mean, which would be engagement-weighted toward members who logged more periods). The fitted model is *sbp*_*c*ℎ*ange_it_* = *β*_0_ + *β*_w,w*it*ℎ*in*_*_it_* + *^β^*w,*bet*w*een_i_* ^+^ *^β^*w*k*,w*eek*_*c_it_* ^+^ *^β^base*,*baseline*_*SBP*_*c_i_* ^+^ *^β^en*6,*n*_*lo*6*in*_*days*_*c_it_* ^+ *u*^*i* ^+^ *^ε^it*^, with the within-^ and between-member terms entered as separate fixed effects and *u_i_* a member-level random effect; *week*_*c* (centred bi-weekly period index) and *baseline*_*SBP*_*c* (centred baseline systolic pressure) are required covariates, as both the weight and systolic series are cumulative from baseline, so a shared program-time trend or regression to the mean would otherwise leak into either term, and n_login_days_c (centred weekly engagement) is included as a further potential confounder of both series. In R version 4.6.1 (R Core Team), the model was specified as y ∼ 1 + within_x + between_x + period_c + baseline_outcome_c + n_login_days_c + (1 + within_x | user_id) and fitted by REML wit using ‘nlme’ version 3.1-169 (53) nlme::lme(fixed = y ∼ …, random = ∼1 + within_x | user_id, correlation = corAR1(form = ∼ period | user_id), method = “REML”). The headline quantity is the Mundlak contrast, *β*_w_ − *β_b_*, with its own confidence interval and p-value computed from this same AR(1)-corrected covariance matrix: a contrast whose interval excludes zero rejects the (implicit) assumption behind the pooled correlation that the two are equal; an interval that includes zero means this design could not separate them at this sample size, not that they are equal. To interpret the Mundlak contrast and contextualize our results in regards to this study statistical power and previous estimates reported in the literature (e.g., (54)) we calculated the minimum-detectable effect (MDE) at 80% power at α= 0.05 two-sided test as *MDE_po_*_w*er*,= 0.05_ = *SE* × (*z*_0.975_ + z_power_).

The application’s AI-based meal rating system scores each logged meal against three dietary-arm axes for coaching prioritization, insufficient fiber, inflammatory potential, and expected insulin response, flagging a meal whenever it carries a penalty on a given axis (Supplementary Methods). The same within/between decomposition above was also fitted with four meal-level metrics derived from these flags in place of weight change: the share of a member’s logged meals flagged on each of the three axes, and the share carrying no penalty on any axis, against both systolic and diastolic change, at the same bi-weekly grain.

## 4 Results

Of the members evaluated for eligibility (n =1,730), 1,127 were excluded for insufficient baseline monitoring (n=795 and n=332 with 1 and 2 distinct days of measurement respectively), 274 had an evaluable follow-up window open but no landmark reading observed in it, leaving 329 members in the analytic cohort. Baseline strata comprised 136 members at goal, 74 at Stage 1 and 119 at Stage 2; 62, 34 and 53 members respectively contributed an evaluable observation at the 6-month landmark (6-month completers: sufficient readings in a 6-month window that had fully elapsed by the data cutoff); a further 42 members had sufficient readings in a 6-month window still open at the cutoff and contribute to the 3-month landmark and to the bi-weekly within/between-member analyses, but not to 6-month landmark estimates. Previous reports recommend discarding the data from the first day of measurements as a valid and adopted approach, because first-day home readings can be systematically elevated (41,55,56). However, this cohort did not show neither elevated systolic nor diastolic day-0 readings (day-0 +2.55 mmHg SBP (95% CI −0.83 to 5.93, p=0.139), +0.16 mmHg DBP (95% CI −2.07 to 2.38, p=0.891)), nor the day-0 occasion count (only 11 of the 329 had day-0 BP readings), differed detectably from later days, so the first calendar day was retained in every window rather than dropped.

Baseline characteristics are shown in Table 1 and a participant flow diagram is provided as Figure 1.

**FIGURE 1.**
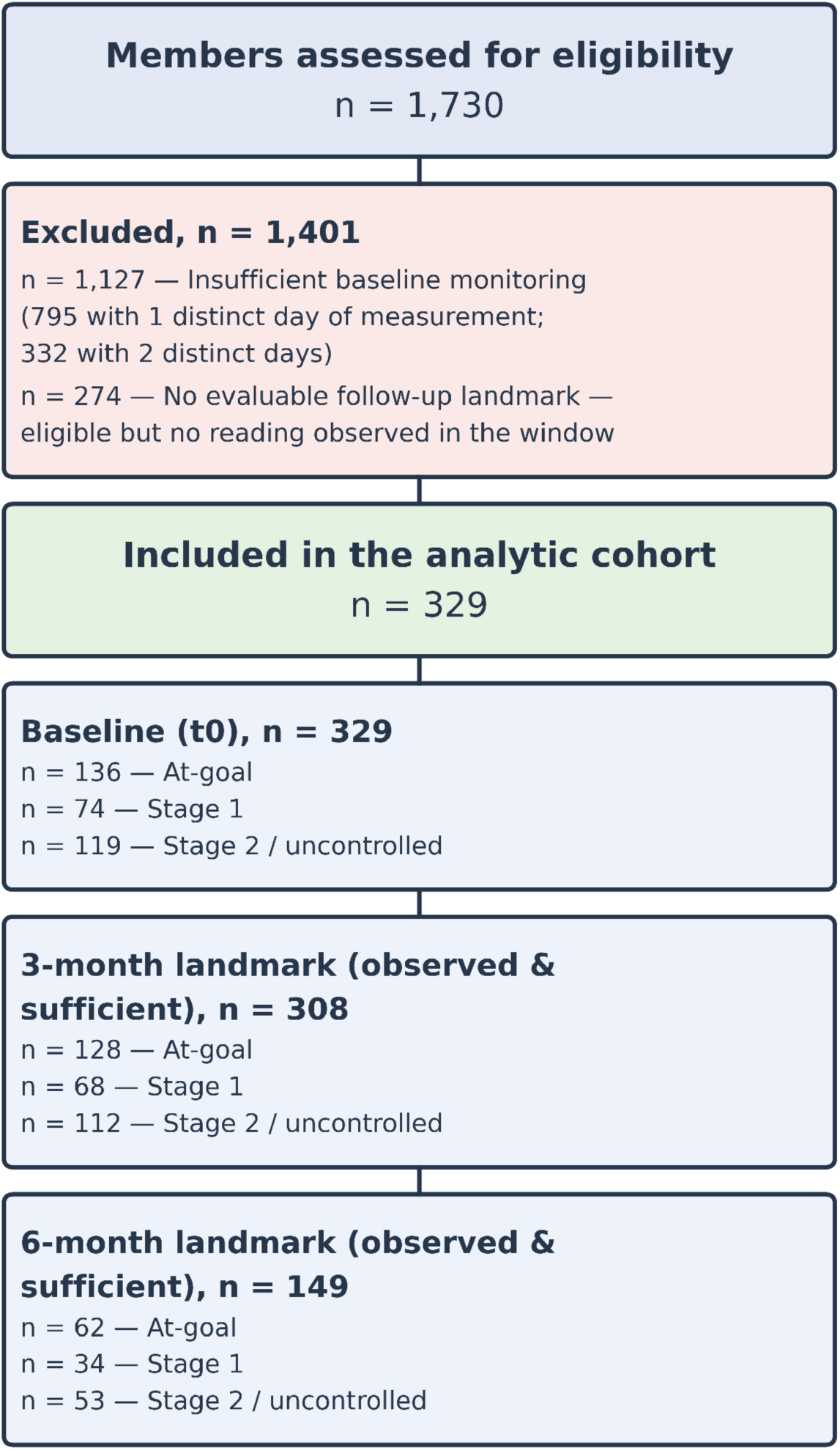
Participant flow. Of the members evaluated for eligibility (n = 1,730), 1,127 were excluded for insufficient baseline monitoring (795 with 1 and 332 with 2 distinct days of measurement) and 274 were excluded for having an evaluable follow-up window open but no landmark reading observed in it, leaving 329 members in the analytic cohort. By baseline home blood pressure: at-goal (n = 136; <130/80 mmHg), Stage 1 (n = 74; 130–134/80–84 mmHg) and Stage 2 (n = 119; ≥135/85 mmHg); 128, 68 and 112 members, respectively, contributed an evaluable observation at the 3-month landmark, and 62, 34 and 53 members, respectively, at the 6-month landmark.

**TABLE 1.**
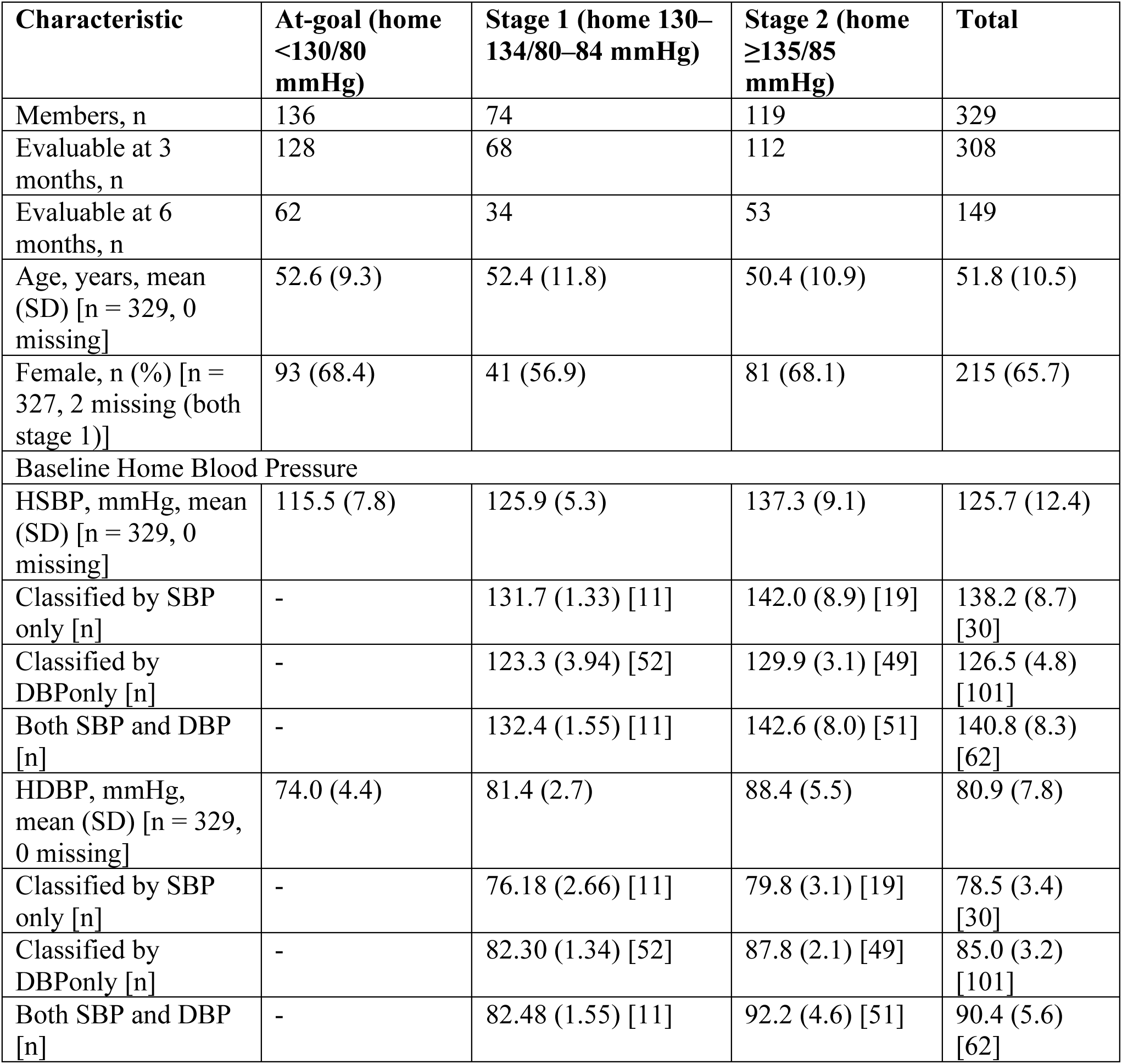
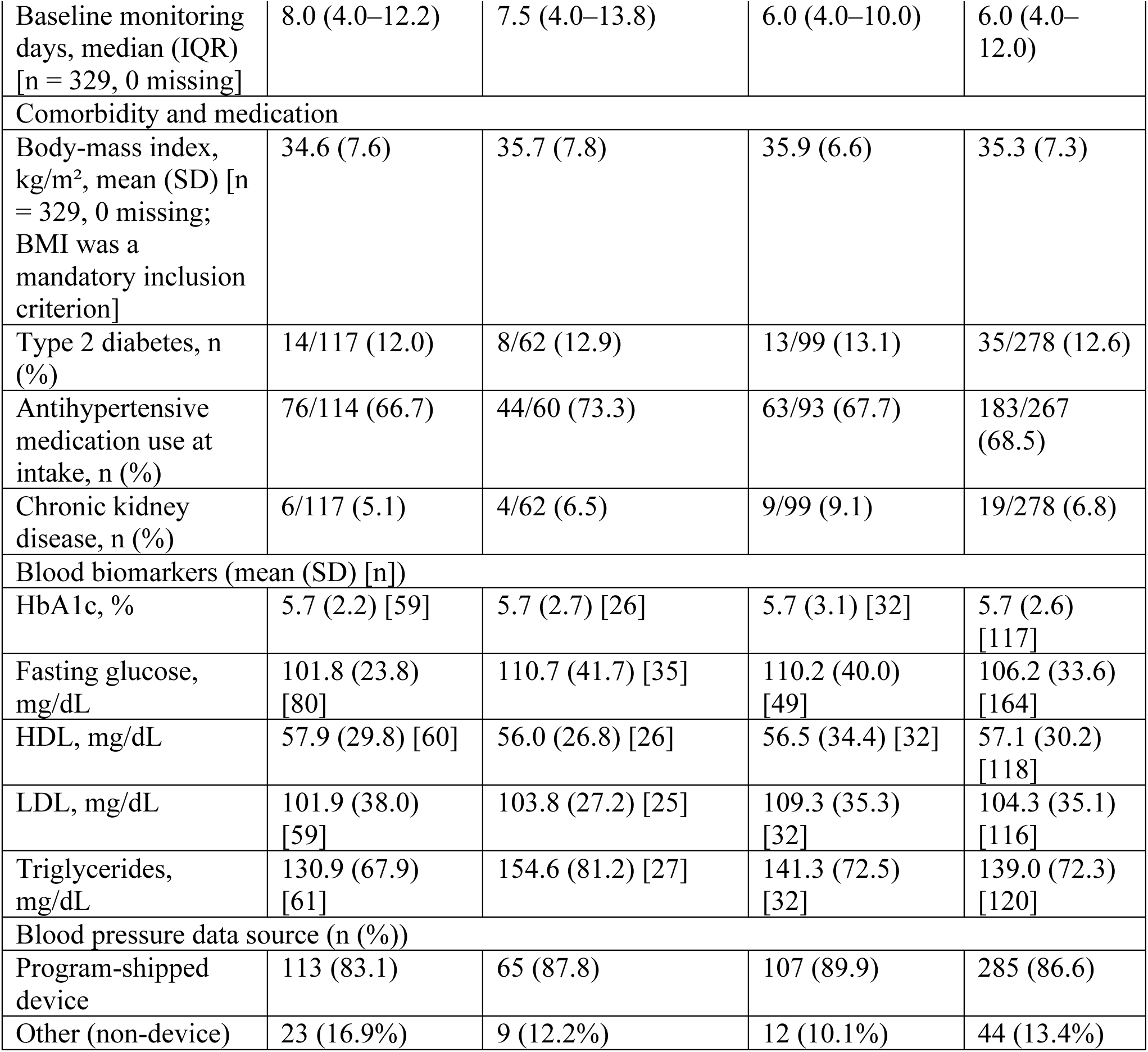
Baseline characteristics by baseline home blood-pressure stratum. Strata: at-goal (<130/80 mmHg), Stage 1 (130–134/80–84), Stage 2 (≥135/85). Continuous variables, mean (SD); categorical, n (%). Biomarker denominators are per-marker and vary independently of stratum, so per-marker n do not sum to the stratum total. HSBP, home systolic blood pressure; HDBP, home diastolic blood pressure; HbA1c, glycated haemoglobin. The baseline hypertension classification is performed as either of the SBP or DBP criteria as per AHA/ACA guidelines (60) and we report the summary statistics for the subgroups classified by either SBP or DBP, or both.

| Characteristic | At-goal (home <130/80 mmHg) | Stage 1 (home 130–134/80–84 mmHg) | Stage 2 (home ≥135/85 mmHg) | Total |
| --- | --- | --- | --- | --- |
| Members, n | 136 | 74 | 119 | 329 |
| Evaluable at 3 months, n | 128 | 68 | 112 | 308 |
| Evaluable at 6 months, n | 62 | 34 | 53 | 149 |
| Age, years, mean (SD) [n = 329, 0 missing] | 52.6 (9.3) | 52.4 (11.8) | 50.4 (10.9) | 51.8 (10.5) |
| Female, n (%) [n = 327, 2 missing (both stage 1)] | 93 (68.4) | 41 (56.9) | 81 (68.1) | 215 (65.7) |
| Baseline Home Blood Pressure |  |  |  |  |
| HSBP, mmHg, mean (SD) [n = 329, 0 missing] | 115.5 (7.8) | 125.9 (5.3) | 137.3 (9.1) | 125.7 (12.4) |
| Classified by SBP only [n] | - | 131.7 (1.33) [11] | 142.0 (8.9) [19] | 138.2 (8.7) [30] |
| Classified by DBP only [n] | - | 123.3 (3.94) [52] | 129.9 (3.1) [49] | 126.5 (4.8) [101] |
| Both SBP and DBP [n] | - | 132.4 (1.55) [11] | 142.6 (8.0) [51] | 140.8 (8.3) [62] |
| HDBP, mmHg, mean (SD) [n = 329, 0 missing] | 74.0 (4.4) | 81.4 (2.7) | 88.4 (5.5) | 80.9 (7.8) |
| Classified by SBP only [n] | - | 76.18 (2.66) [11] | 79.8 (3.1) [19] | 78.5 (3.4) [30] |
| Classified by DBP only [n] | - | 82.30 (1.34) [52] | 87.8 (2.1) [49] | 85.0 (3.2) [101] |
| Both SBP and DBP [n] | - | 82.48 (1.55) [11] | 92.2 (4.6) [51] | 90.4 (5.6) [62] |

| Characteristic | At-goal (home<br><130/80<br>mmHg) | Stage 1 (home 130–<br>134/80–84 mmHg) | Stage 2 (home<br>≥135/85<br>mmHg) | Total |
| --- | --- | --- | --- | --- |
| Baseline monitoring<br>days, median (IQR)<br>[n = 329, 0 missing] | 8.0 (4.0–12.2) | 7.5 (4.0–13.8) | 6.0 (4.0–10.0) | 6.0 (4.0–<br>12.0) |
| Comorbidity and medication |  |  |  |  |
| Body-mass index,<br>kg/m<sup>2</sup>, mean (SD) [n<br>= 329, 0 missing;<br>BMI was a<br>mandatory inclusion<br>criterion] | 34.6 (7.6) | 35.7 (7.8) | 35.9 (6.6) | 35.3 (7.3) |
| Type 2 diabetes, n<br>(%) | 14/117 (12.0) | 8/62 (12.9) | 13/99 (13.1) | 35/278 (12.6) |
| Antihypertensive<br>medication use at<br>intake, n (%) | 76/114 (66.7) | 44/60 (73.3) | 63/93 (67.7) | 183/267<br>(68.5) |
| Chronic kidney<br>disease, n (%) | 6/117 (5.1) | 4/62 (6.5) | 9/99 (9.1) | 19/278 (6.8) |
| Blood biomarkers (mean (SD) [n]) |  |  |  |  |
| HbA1c, % | 5.7 (2.2) [59] | 5.7 (2.7) [26] | 5.7 (3.1) [32] | 5.7 (2.6)<br>[117] |
| Fasting glucose,<br>mg/dL | 101.8 (23.8)<br>[80] | 110.7 (41.7) [35] | 110.2 (40.0)<br>[49] | 106.2 (33.6)<br>[164] |
| HDL, mg/dL | 57.9 (29.8) [60] | 56.0 (26.8) [26] | 56.5 (34.4) [32] | 57.1 (30.2)<br>[118] |
| LDL, mg/dL | 101.9 (38.0)<br>[59] | 103.8 (27.2) [25] | 109.3 (35.3)<br>[32] | 104.3 (35.1)<br>[116] |
| Triglycerides,<br>mg/dL | 130.9 (67.9)<br>[61] | 154.6 (81.2) [27] | 141.3 (72.5)<br>[32] | 139.0 (72.3)<br>[120] |
| Blood pressure data source (n (%)) |  |  |  |  |
| Program-shipped<br>device | 113 (83.1) | 65 (87.8) | 107 (89.9) | 285 (86.6) |
| Other (non-device) | 23 (16.9%) | 9 (12.2%) | 12 (10.1%) | 44 (13.4%) |

### 4.1 Home blood pressure change by stratum and landmark

Among members with Stage-2 hypertension at baseline, unadjusted change was −5.48 mmHg (95% CI −7.51 to −3.45; p = 4.6 × 10⁻⁷) at 3 months and −10.17 mmHg (95% CI −12.94 to −7.40; p = 1.3 × 10⁻⁹) at 6 months, paired with its RTM-corrected change of −1.93 mmHg (95% CI −4.45 to 0.60; p = 0.13) at 3 months and −5.50 mmHg (95% CI −8.50 to −2.50; p = 0.00037) at 6 months (Table 2, Figure 2 and Figure S1). Among members at Stage 1, unadjusted change was −2.48 mmHg (95% CI −4.58 to −0.38; p = 0.022) at 3 months and −4.36 mmHg (95% CI −7.43 to −1.28; p = 0.0069) at 6 months, with RTM-corrected change of −2.07 mmHg (95% CI −4.46 to 0.31; p = 0.088) at 3 months and −3.33 mmHg (95% CI −6.40 to −0.26; p = 0.033) at 6 months (Table 2, Figure 2 and Figure S1). Diastolic blood pressure change results followed the same pattern as systolic, at a smaller absolute magnitude. Among members with Stage-2 hypertension at baseline, unadjusted change was −3.19 mmHg (95% CI −4.48 to −1.90; p = 3.2 × 10⁻⁶) at 3 months and −5.24 mmHg (95% CI −6.95 to −3.54; p = 1.0 × 10⁻⁷) at 6 months, paired with its RTM-corrected change of −1.46 mmHg (95% CI −3.08 to 0.16; p = 0.078) at 3 months and −3.50 mmHg (95% CI −5.42 to −1.59; p = 0.00037) at 6 months (Table 2, Figure 2 and Figure S1). Among members at Stage 1, unadjusted change was −1.76 mmHg (95% CI −3.02 to −0.50; p = 0.0071) at 3 months and −2.02 mmHg (95% CI −3.96 to −0.07; p = 0.042) at 6 months, with RTM-corrected change of −1.37 mmHg (95% CI −2.81 to 0.07; p = 0.062) at 3 months and −1.48 mmHg (95% CI −3.40 to 0.43; p = 0.13) at 6 months (Table 2, Figure 2 and Figure S1). Among members at goal, the internal negative control, home diastolic pressure rose unadjusted by 0.82 mmHg (95% CI −0.21 to 1.85; p = 0.12) at 3 months and 1.16 mmHg (95% CI −0.22 to 2.54; p = 0.097) at 6 months, compared to the RTM-corrected change of −0.18 mmHg (95% CI −1.48 to 1.11; p = 0.78) and +0.30 mmHg (95% CI −1.29 to 1.90; p = 0.71), neither of which excluded zero (Table 2, Figure 2 and Figure S1). Supplementary Table S1 presents summary monitoring density for each Table 2 stratum x landmark cell showing these are similar and comparable across stratum x landmark groups.

**FIGURE 2.**
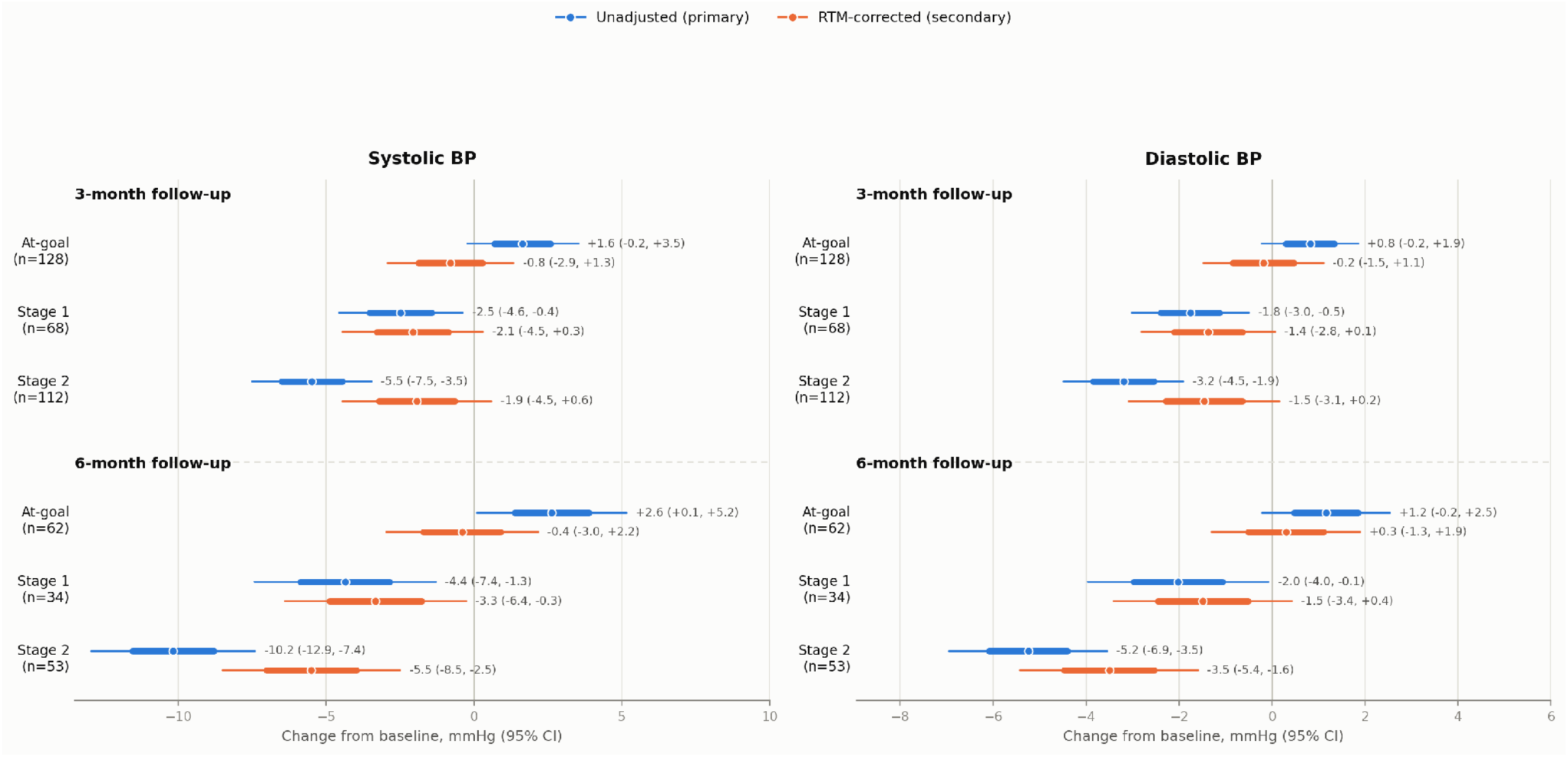
Change in home SBP and DBP estimated at 3 and 6 months landmark stratified by baseline HBP hypertension group (at-goal, Stage 1 and Stage 2) from the unadjusted (colored blue) and RTM-corrected (colored orange) models. SBP and DBP estimated change is presented as mean with ±1 std error (thick line) and 95% CI (thin line) with mean and CI reported as text next to it.

**TABLE 2.** Change in home systolic and diastolic blood pressure from baseline, by baseline stratum and landmark. Unadjusted, mean paired within-member change (Δ mmHg, 95% CI, t-distribution); RTM-adjusted, estimated marginal mean from the reduced ANCOVA linear mixed model (centred baseline blood pressure, stratum × landmark interaction, member-level random intercept), evaluated at the cohort grand-mean baseline. n, members contributing an evaluable observation in that cell.

| Blood pressure and Stratum | Landmark | N participants | Unadjusted $\Delta$ mmHg (95% CI) | p-value | RTM Adjusted $\Delta$ mmHg (95% CI) | p-value |
| --- | --- | --- | --- | --- | --- | --- |
| Systolic Blood Pressure |  |  |  |  |  |  |
| At-goal | 3 mo | 128 | +1.64 (−0.23 to 3.52) | 0.085 | −0.79 (−2.93 to 1.34) | 0.466 |
|  | 6 mo | 62 | +2.63 (0.11 to 5.16) | 0.041 | −0.40 (−2.98 to 2.17) | 0.759 |
| Stage 1 | 3 mo | 68 | −2.48 (−4.58 to −0.38) | 0.022 | −2.07 (−4.46 to 0.31) | 0.088 |
|  | 6 mo | 34 | −4.36 (−7.43 to −1.28) | 0.0069 | −3.33 (−6.40 to −0.26) | 0.033 |
| Stage 2 | 3 mo | 112 | −5.48 (−7.51 to −3.45) | $4.6 \times 10^{-7}$ | −1.93 (−4.45 to 0.60) | 0.134 |
| | 6 mo | 53 | −10.17 (−12.94 to −7.40) | $1.3 \times 10^{-9}$ | −5.50 (−8.50 to −2.50) | $3.7 \times 10^{-4}$ |
| Diastolic Blood Pressure |  |  |  |  |  |  |
| At-goal | 3 mo | 128 | +0.82 (−0.21 to 1.85) | 0.118 | −0.18 (−1.48 to 1.11) | 0.780 |
|  | 6 mo | 62 | +1.16 (−0.22 to 2.54) | 0.097 | +0.30 (−1.29 to 1.90) | 0.711 |
| Stage 1 | 3 mo | 68 | −1.76 (−3.02 to −0.50) | 0.0071 | −1.37 (−2.81 to 0.07) | 0.062 |
|  | 6 mo | 34 | −2.02 (−3.96 to −0.07) | 0.042 | −1.48 (−3.40 to 0.43) | 0.128 |
| Stage 2 | 3 mo | 112 | −3.19 (−4.48 to −1.90) | $3.2 \times 10^{-6}$ | −1.46 (−3.08 to 0.16) | 0.078 |
| | 6 mo | 53 | −5.24 (−6.95 to −3.54) | $1.0 \times 10^{-7}$ | −3.50 (−5.42 to −1.59) | $3.7 \times 10^{-4}$ |

### 4.2 Internal negative control

Among members at goal at baseline (the internal negative control), home systolic pressure rose unadjusted by 1.64 mmHg (95% CI −0.23 to 3.52) at 3 months and 2.63 mmHg (95% CI 0.11 to 5.16) at 6 months, and after RTM correction changed by −0.79 mmHg (95% CI −2.93 to 1.34) and −0.40 mmHg (95% CI −2.98 to 2.17). Diastolic pressure followed a similar pattern, rising unadjusted and reaching −0.18 mmHg (95% CI −1.48 to 1.11) and +0.30 mmHg (95% CI −1.29 to 1.90) after correction (Table 2). The fitted coefficient on centred baseline pressure, a single slope common to all strata, was −0.284 mmHg per mmHg for systolic (95% CI −0.419 to −0.150; p = 4.1 × 10⁻⁵) and −0.192 mmHg per mmHg for diastolic (95% CI −0.328 to −0.057; p = 0.0055).

### 4.3 Weight and systolic pressure change

Across the full cohort, mean weight change from baseline was −7.50 lb (95% CI −8.85 to −6.14, n = 226, p < 0.001) at 3 months and −13.97 lb (95% CI −16.98 to −10.96, n = 124, p < 0.001) at 6 months.

As a secondary, non-confirmatory analysis, we decomposed the weight-change/systolic-change relationship into within-member (bi-weekly, period-to-period) and between-member (member-to-member) components using a Mundlak (57) model analysis which aims to test the possible mediation effect of weight loss on SBP change (Methods). This decomposition was fit on bi-weekly home-monitoring data from this cohort on members with at least one evaluable bi-weekly period of paired weight and home BP readings (715 member-periods, 114 members). Members’ overall weight change across the observation period was associated with their overall SBP change (between-member component: β = 0.44 mmHg per percentage point of baseline body weight lost, SE = 0.12, p < 0.001); a period-to-period, within-member association was not detected (β = 0.09, SE = 0.12, p = 0.461). A formal contrast of the two components (within minus between) reached nominal significance (−0.35, 95% CI −0.68 to −0.02, p = 0.036), indicating the two associations differ from one another at this grain. For DBP the between-member component was β = 0.23 (SE = 0.07, p = 0.002) mmHg per percentage point of baseline body weight lost, the within-member component was β = 0.11 (SE = 0.08, p = 0.142), and the contrast between the two components was −0.12 (95% CI −0.32 to 0.08, p = 0.244); for DBP the two associations did not differ significantly (Table 4). Using the model’s achieved standard error, this cohort’s precision at bi-weekly grain (0.45 mmHg/kg for SBP, 0.28 mmHg/kg for DBP at 80% power, see Methods) is close to a previously reported within-person weight-loss/blood-pressure effect size (∼0.4 mmHg/kg; Morris et al., 2021), supporting the interpretability of the SBP contrast; the DBP result is considered inconclusive.

**TABLE 3.** Attrition-sensitivity estimates by target stratum and landmark, mmHg change vs. baseline. Pattern-mixture (δ=0) is the MNAR delta-adjustment model evaluated at no assumed post-dropout erosion (equivalent to an MAR assumption). Tipping δ is the smallest simulated erosion (mmHg added to each imputed value, swept 0–8) at which the pattern-mixture 95% CI first includes zero, larger is more robust to informative dropout. IPCW is the inverse-probability-of-censoring-weighted estimate, attrition weights fit on age, BMI category, type 2 diabetes, chronic kidney disease, baseline antihypertensive status and device provision, with missing covariates MICE-imputed (m=10, Rubin-pooled). Naive complete-case is the unweighted complete-case ANCOVA. Sample sizes are Stage 1 n=39 (3mo) / 34 (6mo); Stage 2 n=77 (3mo) / 53 (6mo). IPCW’s own weighted effective n runs slightly lower (Stage 1: 28.2/24.5; Stage 2: 69.0/48.4) since it downweights the less-representative completers. All estimates [95% CI], p unadjusted.

| Stratum | Landmark | Pattern-mixture ($\delta=0$) | Tipping $\delta$ | IPCW | Naive complete-case |
| --- | --- | --- | --- | --- | --- |
| Stage 1 | 3mo | −3.48 [−6.18, −0.77], p=.012 | 6 | −4.15 [−7.84, −0.46], p=.029 | −3.76 [−6.55, −0.96], p=.008 |
|  | 6mo | −4.33 [−7.24, −1.42], p=.004 | 6 | −5.46 [−9.38, −1.54], p=.008 | −4.41 [−7.50, −1.31], p=.005 |
| Stage 2 | 3mo | −2.38 [−4.89, 0.12], p=.062 | 0 | −2.91 [−6.52, 0.69], p=.111 | −1.76 [−5.01, 1.49], p=.288 |
|  | 6mo | −4.67 [−7.64, −1.71], p=.002 | 4 | −6.20 [−10.70, −1.71], p=.008 | −6.46 [−10.84, −2.08], p=.004 |

**TABLE 4.** Within/between-person (Mundlak) decomposition of weight change and home blood pressure change, bi-weekly grain.

| Outcome | Members | Periods | Within $\beta$ (SE, p) | Between $\beta$ (SE, p) | Contrast $\beta$ (95% CI, p) |
| --- | --- | --- | --- | --- | --- |
| SBP | 114 | 715 | 0.09 (0.12, .461) | 0.44 (0.12, <.001) | -0.35 (-0.68, -0.02; .036) |
| DBP | 114 | 715 | 0.11 (0.08, .142) | 0.23 (0.07, .002) | -0.12 (-0.32, 0.08; .244) |

Members’ meal-level nutrient density (ND)-score flag rates changed over the same period. The share of logged meals flagged for insufficient fiber declined from baseline at both 3 months (mean paired within-member change −0.106, 95% CI −0.147 to −0.064, n = 131, p < 0.001) and 6 months (−0.138, 95% CI −0.206 to −0.070, n = 68, p < 0.001); the share flagged clean on every scored dimension rose at both landmarks (3 months: +0.032, 95% CI 0.003 to 0.061, p = 0.033; 6 months: +0.103, 95% CI 0.047 to 0.159, p < 0.001). The insulin-associated flag rate declined at 3 months (−0.062, 95% CI −0.112 to −0.011, p = 0.017) but not at 6 months (−0.044, 95% CI −0.115 to 0.027, p = 0.216); the inflammation-associated flag rate did not change significantly at either landmark (3 months: +0.011, 95% CI −0.034 to 0.055, p = 0.639; 6 months: −0.052, 95% CI −0.131 to 0.028, p = 0.201). Fiber and the clean-meal rate therefore show the most consistent evidence of sustained dietary change over the program.

We applied the same within/between decomposition (See Methods) to all four meal-level flag-rate metrics against SBP and DBP change, at the same bi-weekly grain (443 member-periods, 73 members). The between-member component was associated with DBP change for the fiber-flag rate (β = 3.93 mmHg per unit flag-rate, SE = 1.87, p = 0.036) and with SBP change for the clean-meal rate (β = −8.68, SE = 3.99, p = 0.030); the fiber-flag rate’s between-member component was not associated with SBP change (β = 5.47, SE = 3.10, p = 0.078), and the clean-meal rate’s was more weakly associated with DBP change (β = −4.84, SE = 2.52, p = 0.055). The insulin– and inflammation-flag rates showed no association with either outcome (between-member p = 0.40–0.79 across all four models). No within-member component or contrast reached nominal significance in any of the eight models (within: p > 0.08 throughout; contrast: SBP-fiber p = 0.292, DBP-fiber p = 0.179, SBP-clean p = 0.091, DBP-clean p = 0.207; other contrasts p > 0.27) (Table 5). These exploratory, hypothesis-generating comparisons were examined across eight outcome pairs without adjustment for multiple comparisons.

**TABLE 5.** Within/between-person (Mundlak) decomposition of meal-level dietary flag-rate change and home blood pressure change, bi-weekly grain.

| Predictor | Outcome | Members | Periods | Within $\beta$ (SE, p) | Between $\beta$ (SE, p) | Contrast $\beta$ (95% CI, p) |
| --- | --- | --- | --- | --- | --- | --- |
| Fiber-flag rate | SBP | 73 | 443 | 1.55 (2.06, .450) | 5.47 (3.10, .078) | -3.92 (-11.22, 3.38; .292) |
| Fiber-flag rate | DBP | 73 | 443 | 0.81 (1.39, .563) | 3.93 (1.87, .036) | -3.12 (-7.68, 1.43; .179) |
| Inflammation-flag rate | SBP | 73 | 443 | 2.88 (1.68, .087) | 1.85 (3.53, .600) | 1.03 (-6.66, 8.71; .793) |
| Inflammation-flag rate | DBP | 73 | 443 | 0.32 (1.38, .818) | 0.83 (2.13, .695) | -0.52 (-5.49, 4.45; .838) |
| Insulin-flag rate | SBP | 73 | 443 | -1.48 (2.15, .490) | 1.41 (2.77, .611) | -2.89 (-9.62, 3.83; .399) |
| Insulin-flag rate | DBP | 73 | 443 | -0.83 (1.28, .516) | 1.42 (1.68, .399) | -2.25 (-6.30, 1.80; .277) |
| Clean-meal rate | SBP | 73 | 443 | -1.33 (2.33, .566) | -8.68 (3.99, .030) | 7.34 (-1.16, 15.85; .091) |
| Clean-meal rate | DBP | 73 | 443 | -1.31 (1.47, .371) | -4.84 (2.52, .055) | 3.52 (-1.96, 9.00; .207) |

### 4.4 Sensitivity analyses

The results were robust to the sensitivity analyses performed. The subgroup with the greatest impact were the 285 of 329 members (86.6%) with device-provided readings. Restricting the RTM model to those members moved the Stage-2/6-month estimate from −5.50 mmHg in the full sample to −5.91 mmHg (95% CI −9.13 to −2.70; p = 0.00031; n = 50) (Supplementary Table S2).

Adjustment for age, BMI category, T2D, CKD and baseline antihypertensive use was similarly inconsequential. Refitting the adjusted model on a fixed complete-case sample (262 members, 365 observations) with each of age, T2D, CKD and antihypertensive status omitted in turn moved the 6-month Stage 2 estimate (−5.21 mmHg with all five retained) by no more than 0.07 mmHg in either direction (largest excursion 0.07 mmHg omitting age, to −5.28 mmHg; only 0.006 mmHg omitting diabetes, to −5.21 mmHg; Supplementary Tables S3, S4); omitting all five together (equivalent to the RTM model on this same complete-case sample) gave −5.43 mmHg (95% CI −8.84 to −2.01), a total excursion of 0.22 mmHg. In the all-five-covariate model itself, the chronic-kidney-disease term’s own coefficient was +4.55 mmHg (95% CI 0.03 to 9.08; p = 0.049), although it is supported by a small sample size n = 19. Subgroup estimates are reported descriptively by baseline antihypertensive use (−7.21 mmHg, 95% CI −11.27 to −3.16 among users, n = 30; −0.91 mmHg, 95% CI −7.21 to 5.38 among non-users, n = 9, below this analysis’s 10-member interpretability threshold) and by reported T2D (−0.08 mmHg, 95% CI −11.26 to 11.09 among the n = 4 members with diabetes, likewise below threshold; −5.61 mmHg, 95% CI −9.20 to −2.03 among the 35 members without), with the diabetes-positive and non-antihypertensive-user cells below the interpretability threshold for assessing effect modification (Supplementary Table S5).

Three further sensitivity analyses probed structural features of the RTM-corrected model. Adding calendar-month season terms to the reduced model left the SBP correction at 6 months largely unchanged (Stage 1: −3.81 mmHg, 95% CI −6.90 to −0.72; Stage 2: −5.72 mmHg, 95% CI −8.71 to −2.72, versus −3.33 mmHg and −5.50 mmHg without seasonal adjustment), and neither harmonic term was itself associated with SBP change (sine term, p = 0.089; cosine term, p = 0.369). We also tested whether a model allowing the RTM slope to differ by stratum fit better than the pooled single-slope specification, by adding a baseline-SBP × stratum interaction to the reduced model and comparing the two by a likelihood-ratio test on nested maximum-likelihood fits. The interaction term was not significant (χ² = 1.94, df = 2, p = 0.38), consistent with the pooled single-slope specification used in the primary reduced model; under the interaction model itself the Stage-2/6-month estimate was −5.08 mmHg (95% CI −8.56 to −1.61) and the Stage-1/6-month estimate was −3.69 mmHg (95% CI −6.79 to −0.58). An alternative RTM correction, formed from the at-goal stratum’s own baseline-to-change slope (−0.316 mmHg per mmHg, 95% CI −0.535 to −0.097), applied to the other strata by the delta method, gave a similar pattern to the pooled-slope correction at 6 months (Stage 1: −3.96 mmHg, 95% CI −6.94 to −0.98; Stage 2: −5.85 mmHg, 95% CI −9.87 to −1.84, versus −3.33 mmHg and −5.50 mmHg from the pooled-slope model).

Informative attrition did not materially affect the estimates. The pattern-mixture, IPCW, and naive complete-case methods agree in direction at both landmarks in both target strata, and the estimates strengthen from 3 to 6 months. The Stage-1 estimates remain non-null under a simulated missing-not-at-random erosion of up to 5 mmHg, with the pattern-mixture CI first crossing zero at δ = 6 mmHg at both landmarks; the Stage-2/6-month estimate remains non-null to 3 mmHg of erosion (tipping δ = 4 mmHg). The Stage-2/3-month cell had a tipping point at δ = 0, reflecting its non-significant base-case pattern-mixture estimate (p = .062) (Table 3). The estimates obtained at each evaluated δ from 0 to 8 mmHg for SBP at 3 and 6 months are presented for Stage 1 and Stage 2 in Supplementary Tables S6 and S7, respectively.

Comparing the 191 members with sufficient readings in the month-6 window (the 149 6-month completers plus the 42 whose window was still open at the data cutoff) against the 138 without, the two groups were comparable on home-BP stratum, baseline SBP and DBP, 3-month SBP and DBP change, age, gender, BMI category, diabetes, and baseline antihypertensive-medication use (all standardised differences < 0.17; all p > 0.2), but differed materially on BP-measurement modality (standardised difference = 0.367; p = 4.7 × 10⁻⁵) (Supplementary Table S8).

The baseline-window ICC (1) *ρ* was 0.62 (n = 1,917 members with a day-level baseline reading, REML random-intercept fit). At month 6, the classical reliability-based RTM expectation for the change score was −4.80 mmHg at Stage 2, closely consistent with the RTM-corrected secondary model’s own implied change for that cell, −4.67 mmHg (95% CI −8.71 to −0.64). At Stage 1 it was −0.48 mmHg against a model-implied −1.03 mmHg (95% CI −5.28 to 3.23), and at goal it was +3.46 mmHg against a model-implied +3.03 mmHg (95% CI −0.53 to 6.60), the positive sign at goal is expected for an internal negative control. All three strata’s reliability-based and model-implied figures agree in sign and lie within roughly half a point of each other, corroborating the RTM-corrected estimates from a source of information (measurement reliability alone) that shares no fitted parameter with the pooled-slope model producing it.

## 5 Discussion

In a contemporary single-arm cohort of a digital food-as-medicine program, home systolic pressure among members with predominantly treated, HBP Stage 2 hypertension at baseline was associated with an unadjusted decline of 10.17 mmHg at 6 months (95% CI −12.94 to −7.40; p = 1.3 × 10⁻⁹) and a RTM-corrected decline of 5.50 mmHg (95% CI −8.50 to −2.50; p = 0.00037) (Table 2 and Figure 2). Approximately 45.9% (95% CI 13.2–78.7%) of the unadjusted decline was attributable to regression toward the baseline mean.

The SBP reductions reported in this study are comparable in magnitude and direction to previous reports. Our unadjusted figure is of the same order as the Stage 2 digital and food-as-medicine cohorts, which report mean SBP reductions ranging from −8.1 mmHg to −14.2 mmHg in comparably elevated subgroups (4–7).

Previous reports with designs that hold a concurrent comparator or apply their own baseline correction report smaller net effects, closer to our RTM-corrected. These comparator-arm estimates span from −2.76 mmHg (95% CI −3.94 to −1.58) to −11.4 mmHg (95% CI −15.9 to −6.9) (8–11). Our RTM-corrected SBP change estimate of 5.50 mmHg falls within this range, indicating that RTM correction brings single-arm estimates into line with those obtained from comparator-arm designs. As hypothesized in our design, the at-goal stratum behaved as an internal negative control. Because this stratum is defined internally rather than as a formal negative control in the sense of Lipsitch et al. (18), we tested and provided evidence of the group’s BP change trajectory. We hypothesized that members already at goal had little headroom to improve, so their unadjusted rise, 2.63 mmHg systolic (95% CI 0.11 to 5.16) and 1.16 mmHg diastolic (95% CI −0.22 to 2.54) at 6 months, is the expected signature and aligned with regression to the mean rather than of deterioration under the program. Both rises disappeared once baseline pressure was accounted for (−0.40 mmHg systolic, 95% CI −2.98 to 2.17; +0.30 mmHg diastolic, 95% CI −1.29 to 1.90), and the fitted slope on centred baseline pressure was negative and clearly non-null for both systolic (−0.284 mmHg per mmHg, 95% CI −0.419 to −0.150) and diastolic pressure (−0.192 mmHg per mmHg, 95% CI −0.328 to −0.057). The correction therefore removes baseline-dependent movement in the one stratum in which such movement can only be regression to the mean. Because the strata are themselves defined by baseline pressure, the fitted baseline slope could in principle combine regression to the mean with any genuine baseline-dependent difference in response. A likelihood-ratio test comparing the pooled slope to one allowing it to vary by stratum found no evidence of stratum-specific slopes (χ² = 1.94, df = 2, p = 0.38), arguing against the most direct alternative.

Diastolic blood pressure change was directionally concordant with systolic at a smaller absolute magnitude. Among members at Stage 2 at baseline, unadjusted diastolic pressure fell by 5.24 mmHg at 6 months (95% CI −6.95 to −3.54), and the RTM-corrected estimate of −3.50 mmHg (95% CI −5.42 to −1.59) excluded zero, whereas the 3-month estimate (−1.46 mmHg, 95% CI −3.08 to 0.16) did not. The concordant direction across the two pressures supports the internal coherence of the SBP finding.

Our results indicate the main findings for SBP are robust to the potential confounders evaluated. Measurement modality did not drive the finding since restricting the model to members who received a program-shipped cellular cuff moved the 6-month Stage 2 estimate by 0.41 mmHg (from −5.50 to −5.91 mmHg), and entering modality as a fixed effect moved it by 0.10 mmHg. Because 86.6% of the cohort had device-provided readings, modality-specific variation in effect magnitude was not examined further. Similarly, age, BMI category, self-reported T2D, CKD and baseline antihypertensive use were likewise not material, with a total excursion of 0.22 mmHg across specifications and substantially overlapping estimates within and outside each subgroup (Supplementary Tables S3–S5). Similarly, the attrition analyses indicate the results are largely insensitive to the evaluated confounding effect of missing-non-at-random and the results indicate the results are more robust from 3 to 6 months landmarks (Table 3).

The corrected 6-month decline of 5.50 mmHg among members at Stage 2 is of a magnitude with potential population-level relevance, since reductions of this order in systolic pressure are associated with materially lower incidence of stroke and major cardiovascular events in meta-analyses of blood pressure lowering treatments (1,2). As baseline staging here is based on home screening rather than clinical diagnosis, this comparison is offered as context for the clinical relevance of the observed change rather than as a direct estimate of cardiovascular risk reduction in this cohort.

As a mechanistic exploration, we tested whether members’ weight change and systolic change shared a common within– and between-member relationship. This within/between-person (57) decomposition found the between-member component nominally significant (β = 0.44 mmHg per percentage point of baseline weight lost, p < 0.001) while the within-member, period-to-period component was not (β = 0.09, p = 0.461); a formal contrast of the two reached nominal significance (p = 0.036), indicating the two associations differ from one another. Converting this cohort’s own units (percentage of baseline weight) to kilograms using its mean analytic-sample baseline weight (103.3 kg), this design’s achieved precision (a minimum detectable Mundlak contrast of 0.45 mmHg/kg for SBP and 0.28 mmHg/kg for DBP at 80% power) is close to previously reported community weight-loss cohorts (∼0.4 mmHg/kg; Morris et al., 2021) and comfortably below the larger ecological meta-regression estimate across randomized weight-loss trials (∼1.05 mmHg/kg, a group-level, not member-level, comparator; Neter et al., 2003). The between-member coefficient itself (0.43 mmHg/kg, p < 0.001) is of a similar order to, though smaller than, Neter’s ecological figure; the within-member point estimate (0.09 mmHg/kg) is well below Morris’s community-cohort figure, so the within-member result is considered inconclusive at this precision. Both within– and between-person coefficients may be influenced by time-varying factors, such as changes in antihypertensive medication during follow-up (58).

Weight loss is therefore unlikely to be the sole pathway, and the program is not designed to act through one. It applies no hypertension-specific dietary protocol; instead it moves several components that independently carry blood-pressure-lowering evidence, reduced alcohol intake, increased dietary fiber and vegetable diversity, reduced ultra-processed food intake, and structured weight loss within a DPP-based curriculum with additional personalization to lifestyle, genetics and microbiome profiles. The observed decline is plausibly the joint product of these, delivered as an integrated bundle; apportioning it among components will require longitudinal dietary and behavioural measures to test mediation (see limitations below). Attribution to any single component would require either component-level exposure data or a factorial design. Importantly, the magnitude of the corrected decline is consistent with what the component literature would imply for a program of this composition.

Consistent with this, members’ meal-level dietary behavior changed unevenly across ND-score domains: the fiber-flag rate and the clean-meal rate both improved at 3 and 6 months, while the insulin-flag rate improved only transiently and the inflammation-flag rate did not change. An exploratory decomposition of these same four flag-rate metrics against blood pressure change, at the same bi-weekly grain as the weight analysis above, found the fiber-flag rate’s between-member component associated with diastolic change (p = 0.036) and the clean-meal rate’s associated with systolic change (p = 0.030, cluster-robust cross-check p = 0.035); the insulin– and inflammation-flag rates showed no association with either outcome (Table 5). This pattern, an association confined to the two metrics that themselves showed sustained behavioral change, is consistent with the fiber component of the program’s dietary guidance specifically, rather than the program overall, and, as exploratory comparisons drawn post-hoc across eight related outcome pairs, provide supporting evidence for fiber’s place among the several independently-supported components named above. This study has limitations. First, the study is single-arm with no concurrent control, so residual secular and seasonal trend is not removable by adjustment for baseline blood pressure. Second, as also reported by others (14), regression to the mean is large in home monitoring: approximately 45.9% (95% CI 13.2–78.7%) of the unadjusted Stage-2 6-month decline is attributable to it by the model-estimated share reported above which we estimated using two complementary approaches, adjusted for, not eliminated. Third, home staging is a screening rather than a diagnostic classification, with reported agreement against ambulatory monitoring in the range κ ≈ 0.2–0.6 and specificity near 53% (59). Fourth, the cohort consists of self-selected, engaged home monitors within a commercial program, which limits generalizability. Fifth, medications are captured at intake only, so change in therapy during follow-up is unobserved and treatment intensification cannot be ruled out as a confounder. Sixth, the internally defined negative control is a majority-medicated, currently controlled stratum rather than an untreated population. This is consistent with its role as an internal negative-control group, since the argument requires only that it be unable to benefit from the intervention, but it limits generalizability of our results. Seventh, lack of longitudinal antihypertensive therapy, sodium intake, alcohol consumption, physical activity or sleep preclude us from testing their possible mediation impact in home BP readings (58). Finally, the lack of contemporaneous non-enrolled series, there is no external comparator against which secular and seasonal trend could be probed by a difference-in-differences or negative-control-outcome analysis. Our results warrant further research, including the possibility to construct a concurrent comparator group, validating home staging against ambulatory or in-clinic measurement in a subsample would establish how the screening classification maps onto diagnostic categories in this population. Capturing medication changes during follow-up, rather than at intake only, would allow treatment intensification to be modelled.

## 6 Conclusion

In a contemporary single-arm cohort of a digital food-as-medicine program, participation was associated with a statistically significant unadjusted decline in home systolic blood pressure of 10.17 mmHg among members with predominantly treated, uncontrolled Stage 2 hypertension at baseline, and with a smaller RTM-corrected decline of 5.50 mmHg, a gap of which approximately 45.9% is attributable to regression toward the baseline mean. Among members with Stage 1 hypertension at baseline, the unadjusted decline was 4.36 mmHg, paired with a smaller RTM-corrected decline of 3.33 mmHg, of which approximately 24% is attributable to regression toward the baseline mean. Exploratory within/between-person analyses linking weight change and meal-level dietary flag rates to blood pressure change point to a plausible behavioral pathway that merits further investigation. A study with a concurrent control is the natural next step to establish causality.

## 7 Data Availability Statement

The data analysed in this study are member-level records from a commercial digital health program and cannot be shared publicly due to consent restrictions.

## 8 Code Availability Statement

The custom analysis code used to generate the results reported here is available from the corresponding author upon reasonable request.

## 9 Ethics Statement

The Institutional Review Board of E&I Review Services reviewed and approved the study protocol (protocol code #18053, approved 05/22/2018). Before being included in the study, all participants provided electronic informed consent as part of their written consent process.

## 10 Author Contributions

IP: Conceptualization, Data curation, Formal analysis, Investigation, Methodology, Resources, Software, Validation, Visualization, Writing – original draft, Writing – review & editing.

SVK: Writing – review & editing.

SKS: Data Collection, Methodology, Writing – review & editing.

GS: Writing review & editing.

DEA: Conceptualization, Writing review & editing.

RS: Conceptualization, Funding acquisition, Project administration, Supervision, Writing – review & editing.

## 11 Funding

The author(s) declare that financial support was received for the research, authorship, and/or publication of this article. This study was funded by Digbi Health, Mountain View, CA, United States.

## 12 Conflict of Interest

All authors are employees and/or contractors of Digbi Health and may hold stocks or stock options on Digbi Health. The digital therapeutics program provided to study participants in this work is a commercially available program developed and marketed by Digbi Health. RS was CEO and founder of Digbi Health. DA has received royalties from Kura Biotech. SK, IP, RS, and DA had patents pending concerning this work. The former conflicts of interest do not alter our adherence to policies on sharing data and materials.

## 13 Supplementary Material

Completed STROBE (cohort-study) and RECORD (routinely-collected-data extension) reporting checklists are provided as Supplementary Material, cited from Methods by reference to the formal STROBE/RECORD papers rather than reproduced in the main text.

In addition, the following Supplementary Material is provided:

Supplementary Figure S1: Home SBP and DBP estimated change at 3 and 6 months by baseline home-BP stratum, unadjusted versus RTM-corrected.

Supplementary Table S1: Monitoring intensity by baseline home-BP stratum and landmark.

Supplementary Table S2: Home SBP change by blood-pressure measurement modality (device-provided versus self-reported).

Supplementary Table S3: Home SBP change by covariate specification, drop-one sensitivity on the complete-case sample.

Supplementary Table S4: Fixed effects of the fully adjusted complete-case model underlying Supplementary Table S3.

Supplementary Table S5: Subgroup refits of the RTM model (antihypertensive use, type 2 diabetes), complete-case sample.

Supplementary Tables S6-S7: Pattern-mixture attrition sensitivity, full erosion-delta curve underlying the tipping-point figure reported in Table 3.

Supplementary Table S8: Completers-versus-non-completers comparability, attrition evidence.

## Supporting information

RECORD checklist

STROBE checklist

## 17 Supplementary Materials

**SUPPLEMENTARY FIGURE S1.**
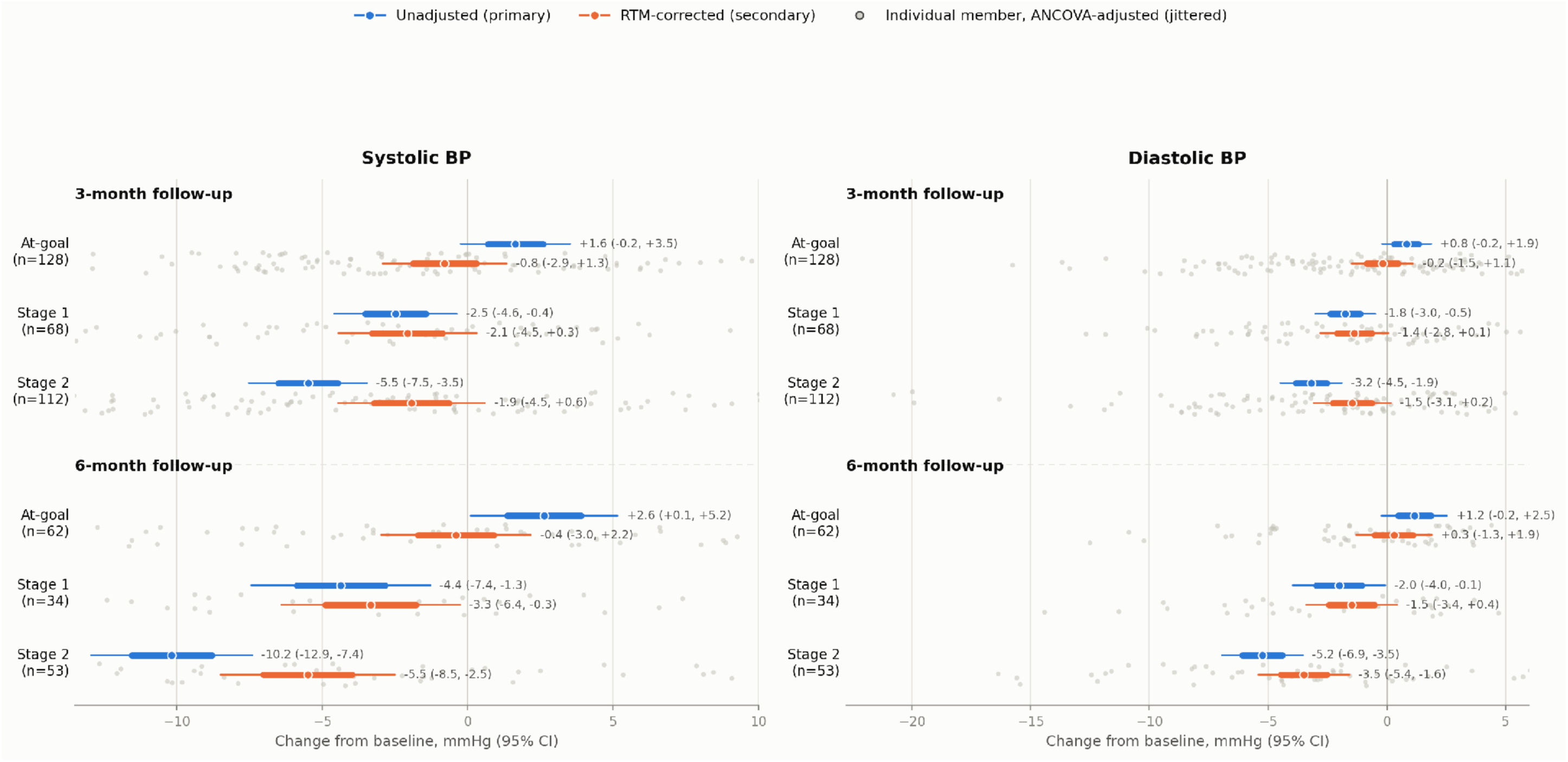
Home SBP and DBP estimated change at 3 and 6 months landmark stratified by baseline HTN group (at-goal, Stage 1 and Stage 2) from the unadjusted (colored blue) and RTM-corrected models (colored orange). Each landmark x HTN group entry includes jittered data points under RTM-corrected results representing the per-member ANCOVA-adjusted change (raw delta minus the reduced model’s pooled baseline-slope × that member’s centered baseline). SBP and DBP estimated change is presented as mean with 1 std error (thick line) and 95% CI (thin line) with mean and CI reported as text next to it.

**SUPPLEMENTARY TABLE S1.** Monitoring intensity by baseline home blood-pressure stratum and landmark. Values are, per stratum × landmark cell: n (members contributing), median day (from baseline) of the last blood-pressure reading used for that landmark, median number of days with at least one reading (monitoring days), median blood-pressure measurement occasions per day, and median readings per occasion.

| Stratum | Landmark | n | Median day of last reading | Median monitoring days | Occasions/day | Readings/occasion |
| --- | --- | --- | --- | --- | --- | --- |
| At-goal | 3mo | 128 | 109.5 | 4.0 | 1.0 | 1.05 |
|  | 6mo | 62 | 196.0 | 3.0 | 1.0 | 1.13 |
| Stage 1 | 3mo | 68 | 111.0 | 5.0 | 1.0 | 1.04 |
|  | 6mo | 34 | 204.5 | 5.5 | 1.0 | 1.18 |
| Stage 2 | 3mo | 112 | 108.0 | 5.0 | 1.0 | 1.02 |
|  | 6mo | 53 | 203.0 | 4.0 | 1.0 | 1.00 |

**SUPPLEMENTARY TABLE S2.**
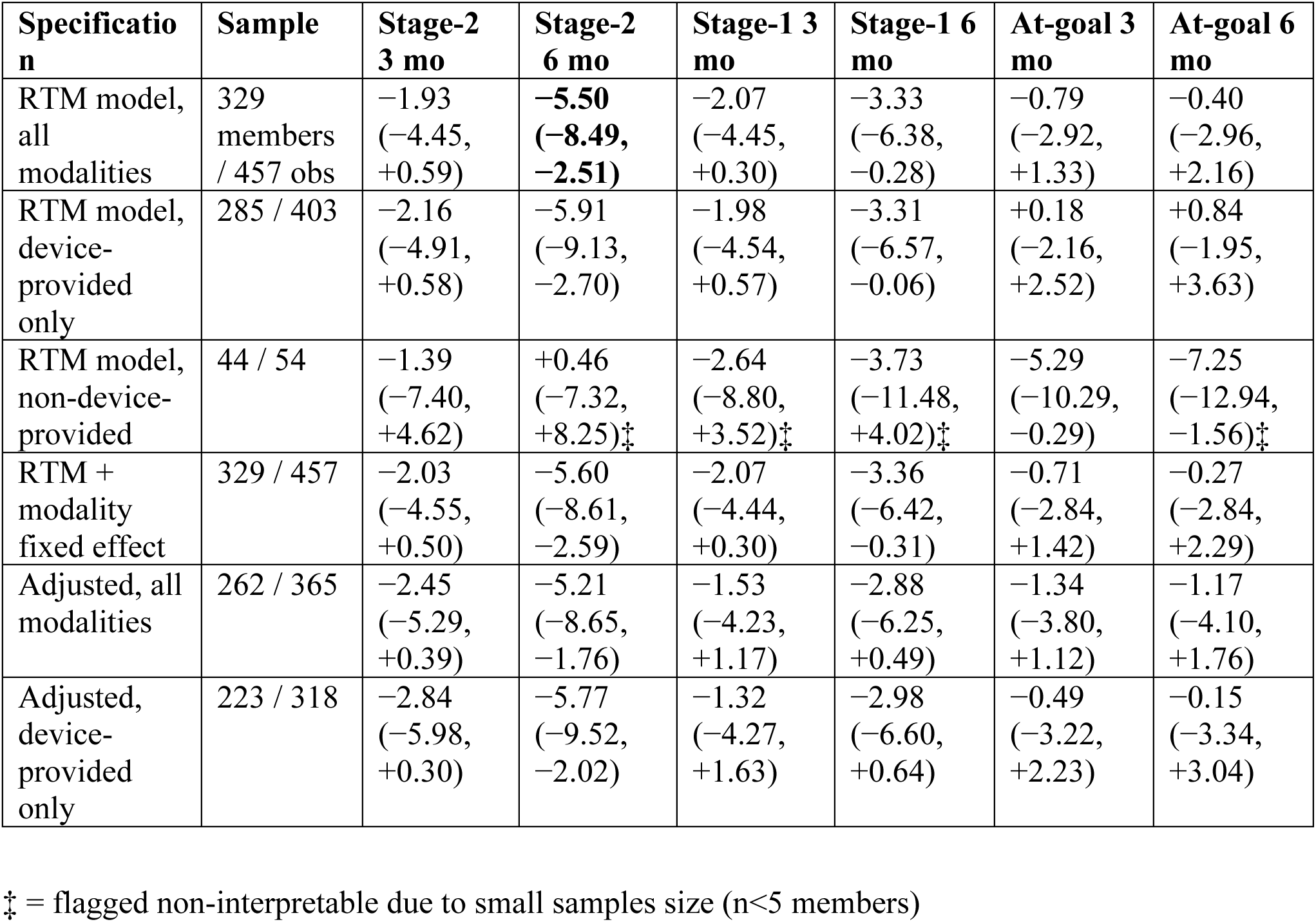
Home systolic blood-pressure changes by measurement modality. Estimated marginal means (mmHg, 95% CI) from design-row linear combinations at each cell’s covariate composition, centred baseline SBP = 0. Adjusted specification: RTM model + age (centred), BMI category, type 2 diabetes, chronic kidney disease, baseline antihypertensive status; complete-case, no imputation. Modality assigned at member level (any dispatched cuff = device-provided). ‡ = flagged non-interpretable due to small samples size (n<5 members)

| Specification | Sample | Stage-2<br>3 mo | Stage-2<br>6 mo | Stage-1 3<br>mo | Stage-1 6<br>mo | At-goal 3<br>mo | At-goal 6<br>mo |
| --- | --- | --- | --- | --- | --- | --- | --- |
| RTM model, all modalities | 329 members / 457 obs | -1.93<br>(-4.45, +0.59) | **-5.50**<br>**(-8.49, -2.51)** | -2.07<br>(-4.45, +0.30) | -3.33<br>(-6.38, -0.28) | -0.79<br>(-2.92, +1.33) | -0.40<br>(-2.96, +2.16) |
| RTM model, device-provided only | 285 / 403 | -2.16<br>(-4.91, +0.58) | -5.91<br>(-9.13, -2.70) | -1.98<br>(-4.54, +0.57) | -3.31<br>(-6.57, -0.06) | +0.18<br>(-2.16, +2.52) | +0.84<br>(-1.95, +3.63) |
| RTM model, non-device-provided | 44 / 54 | -1.39<br>(-7.40, +4.62) | +0.46<br>(-7.32, +8.25)‡ | -2.64<br>(-8.80, +3.52)‡ | -3.73<br>(-11.48, +4.02)‡ | -5.29<br>(-10.29, -0.29) | -7.25<br>(-12.94, -1.56)‡ |
| RTM + modality fixed effect | 329 / 457 | -2.03<br>(-4.55, +0.50) | -5.60<br>(-8.61, -2.59) | -2.07<br>(-4.44, +0.30) | -3.36<br>(-6.42, -0.31) | -0.71<br>(-2.84, +1.42) | -0.27<br>(-2.84, +2.29) |
| Adjusted, all modalities | 262 / 365 | -2.45<br>(-5.29, +0.39) | -5.21<br>(-8.65, -1.76) | -1.53<br>(-4.23, +1.17) | -2.88<br>(-6.25, +0.49) | -1.34<br>(-3.80, +1.12) | -1.17<br>(-4.10, +1.76) |
| Adjusted, device-provided only | 223 / 318 | -2.84<br>(-5.98, +0.30) | -5.77<br>(-9.52, -2.02) | -1.32<br>(-4.27, +1.63) | -2.98<br>(-6.60, +0.64) | -0.49<br>(-3.22, +2.23) | -0.15<br>(-3.34, +3.04) |

‡ = flagged non-interpretable due to small samples size (n<5 members)

**SUPPLEMENTARY TABLE S3.** Home systolic blood-pressure change by covariate specification. Estimated marginal means (mmHg, 95% CI); fixed complete-case sample, 262 members, 365 observations. All rows retain centred baseline SBP, landmark × stratum interaction and member-level random intercept; the “Adjusted” and single-covariate-omission rows additionally retain BMI category, which is dropped along with the other four covariates in the “Omitting all five” and both RTM rows. Excursion at the 6-month uncontrolled cell: 0.07 mmHg (largest single covariate, age); 0.22 mmHg (all five omitted).

| Specification | Uncontrolled 3<br>mo | Uncontrolled 6<br>mo | Stage-1 6 mo | At-goal 6 mo |
| --- | --- | --- | --- | --- |
| Adjusted (all five covariates) | −2.45 (−5.29, +0.39) | −5.21 (−8.65, −1.76) | −2.88 (−6.25, +0.49) | −1.17 (−4.10, +1.76) |
| Omitting age | −2.51 (−5.34, +0.31) | −5.28 (−8.71, −1.85) | −2.87 (−6.24, +0.50) | −1.14 (−4.06, +1.79) |
| Omitting type 2 diabetes | −2.46 (−5.28, +0.37) | −5.21 (−8.65, −1.78) | −2.88 (−6.25, +0.48) | −1.17 (−4.09, +1.76) |
| Omitting chronic kidney disease | −2.55 (−5.40, +0.30) | −5.24 (−8.70, −1.78) | −2.98 (−6.36, +0.41) | −1.13 (−4.07, +1.81) |
| Omitting antihypertensive status | −2.46 (−5.29, +0.38) | −5.25 (−8.68, −1.81) | −2.88 (−6.25, +0.49) | −1.17 (−4.10, +1.76) |
| Omitting all five | −2.67 (−5.47, +0.13) | −5.43 (−8.84, −2.01) | −2.96 (−6.32, +0.41) | −0.99 (−3.90, +1.92) |
| RTM model (no covariates), full sample | −1.93 (−4.45, +0.59) | −5.50 (−8.49, −2.51) | −3.33 (−6.38, −0.28) | −0.40 (−2.96, +2.16) |
| RTM model (no covariates), same complete-case sample | −2.67 (−5.47, +0.13) | −5.43 (−8.84, −2.01) | −2.96 (−6.32, +0.41) | −0.99 (−3.90, +1.92) |

**SUPPLEMENTARY TABLE S4.** Fixed effects of the fully adjusted complete-case model (Supplementary Table S3). Coefficients in mmHg with 95% CI and p-values; random-intercept variance in mmHg². Chronic kidney disease prevalence 7.3% of the 262 complete-case members.

| **Term** | **Coefficient** | **95% CI** | **p** |
| --- | --- | --- | --- |
| Intercept | −0.058 | −6.50, +6.38 | 0.986 |
| Landmark 6 | +0.168 | −2.24, +2.57 | 0.891 |
| Stratum: Stage-1 | −0.290 | −4.04, +3.46 | 0.879 |
| Stratum: uncontrolled | −1.229 | −5.79, +3.33 | 0.597 |
| BMI: obese | −1.923 | −8.05, +4.20 | 0.538 |
| BMI: overweight | −2.497 | −8.84, +3.84 | 0.440 |
| Landmark 6 × Stage-1 | −1.554 | −5.63, +2.53 | 0.455 |
| Landmark 6 ×<br>uncontrolled | −2.886 | −6.57, +0.80 | 0.125 |
| Centred baseline SBP<br>(per mmHg) | −0.321 | −0.48, −0.16 | <0.001 |
| Age (per year,<br>centred) | +0.036 | −0.10, +0.18 | 0.613 |
| Type 2 diabetes | +0.041 | −3.53, +3.62 | 0.982 |
| Chronic kidney<br>disease | +4.551 | +0.03, +9.08 | 0.049 |
| Baseline<br>antihypertensive use | +0.606 | −1.95, +3.16 | 0.642 |
| Random-intercept<br>variance | 62.127 | — | — |

**SUPPLEMENTARY TABLE S5.** Subgroup refits of the RTM model, complete-case sample, home systolic blood pressure. Estimated marginal means (mmHg, 95% CI); descriptive, not a test of interaction. ‡ Cell with fewer than 10 members (this analysis’s interpretability threshold); estimate not interpretable.

| Subgroup | Members | Uncontrolled 3 mo | Uncontrolled 6 mo (n) | Stage-1 3 mo | Stage-1 6 mo | At-goal 3 mo | At-goal 6 mo |
| --- | --- | --- | --- | --- | --- | --- | --- |
| On antihypertensive at baseline | 181 | -4.38<br>(-7.81, -0.95) | -7.21<br>(-11.27, -3.16),<br>n=30 | -0.94<br>(-4.21, +2.32) | -3.83<br>(-7.88, +0.21) | +0.44<br>(-2.52, +3.39) | +1.06<br>(-2.48, +4.60) |
| Not on antihypertensive | 81 | +1.55<br>(-3.24, +6.34) | -0.91<br>(-7.21, +5.38)‡,<br>n=9 | -3.22<br>(-7.94, +1.51) | -1.03<br>(-7.00, +4.95)‡ | -5.15<br>(-9.37, -0.94) | -5.96<br>(-11.01, -0.92) |
| Type 2 diabetes reported | 34 | -3.81<br>(-12.27, +4.66) | -0.08<br>(-11.26, +11.09)‡,<br>n=4 | +4.02<br>(-4.43, +12.47)<br>‡ | -3.00<br>(-11.91, +5.91)‡ | -3.32<br>(-9.75, +3.12) | -2.31<br>(-9.44, +4.81)‡ |
| No type 2 diabetes reported | 228 | -2.11<br>(-5.09, +0.88) | -5.61<br>(-9.20, -2.03),<br>n=35 | -2.31<br>(-5.14, +0.51) | -2.63<br>(-6.27, +1.01) | -1.33<br>(-4.01, +1.34) | -1.29<br>(-4.52, +1.95) |
| Chronic kidney disease reported ‡ | 19 | -1.29<br>(-8.57, +6.00)‡ | -2.53<br>(-11.44, +6.38)‡,<br>n=3 | +5.95<br>(-3.12, +15.03)<br>‡ | +4.62<br>(-4.86, +14.11)<br>‡ | +1.30<br>(-6.56, +9.16)<br>‡ | +6.16<br>(-2.67, +14.99)<br>‡ |
| No chronic kidney disease reported | 243 | -2.41<br>(-5.45, +0.63) | -5.32<br>(-8.96, -1.69),<br>n=36 | -2.11<br>(-4.92, +0.70) | -3.43<br>(-7.01, +0.15) | -1.78<br>(-4.39, +0.82) | -1.79<br>(-4.90, +1.31) |

**SUPPLEMENTARY TABLE S6.**
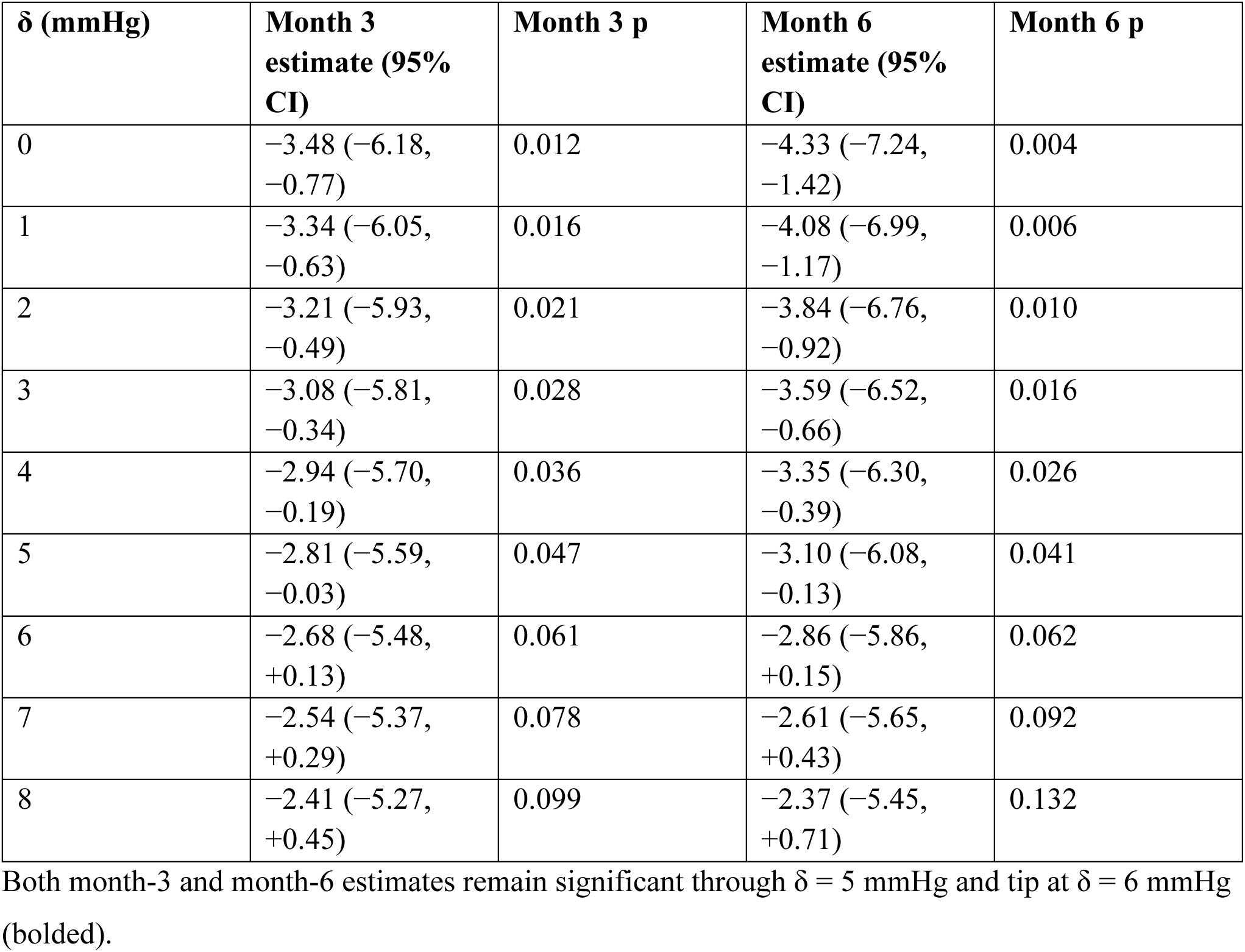
Values are the pattern-mixture model-based mean change in systolic blood pressure from baseline (mmHg, 95% CI) and its raw, unadjusted p-value, re-estimated at each erosion delta δ (0–8 mmHg) swept against every imputed (non-observed) follow-up value; δ = 0 is the base case, assuming missing-at-random. Larger δ represents a progressively larger assumed departure from missing-at-random, applied only in the direction that erodes the observed decline toward the null. The bolded row marks the tipping δ (the smallest value in the grid at which the 95% CI first includes zero), reported as a single figure in Table 3; this table shows the full erosion curve underlying it. CI, confidence interval; δ, erosion delta.

| $\delta$ (mmHg) | **Month 3**<br>**estimate (95%**<br>**CI)** | **Month 3 p** | **Month 6**<br>**estimate (95%**<br>**CI)** | **Month 6 p** |
| --- | --- | --- | --- | --- |
| 0 | –3.48 (–6.18,<br>–0.77) | 0.012 | –4.33 (–7.24,<br>–1.42) | 0.004 |
| 1 | –3.34 (–6.05,<br>–0.63) | 0.016 | –4.08 (–6.99,<br>–1.17) | 0.006 |
| 2 | –3.21 (–5.93,<br>–0.49) | 0.021 | –3.84 (–6.76,<br>–0.92) | 0.010 |
| 3 | –3.08 (–5.81,<br>–0.34) | 0.028 | –3.59 (–6.52,<br>–0.66) | 0.016 |
| 4 | –2.94 (–5.70,<br>–0.19) | 0.036 | –3.35 (–6.30,<br>–0.39) | 0.026 |
| 5 | –2.81 (–5.59,<br>–0.03) | 0.047 | –3.10 (–6.08,<br>–0.13) | 0.041 |
| 6 | –2.68 (–5.48,<br>+0.13) | 0.061 | –2.86 (–5.86,<br>+0.15) | 0.062 |
| 7 | –2.54 (–5.37,<br>+0.29) | 0.078 | –2.61 (–5.65,<br>+0.43) | 0.092 |
| 8 | –2.41 (–5.27,<br>+0.45) | 0.099 | –2.37 (–5.45,<br>+0.71) | 0.132 |

Both month-3 and month-6 estimates remain significant through $\delta = 5$ mmHg and tip at $\delta = 6$ mmHg (bolded).

**SUPPLEMENTARY TABLE S7.**
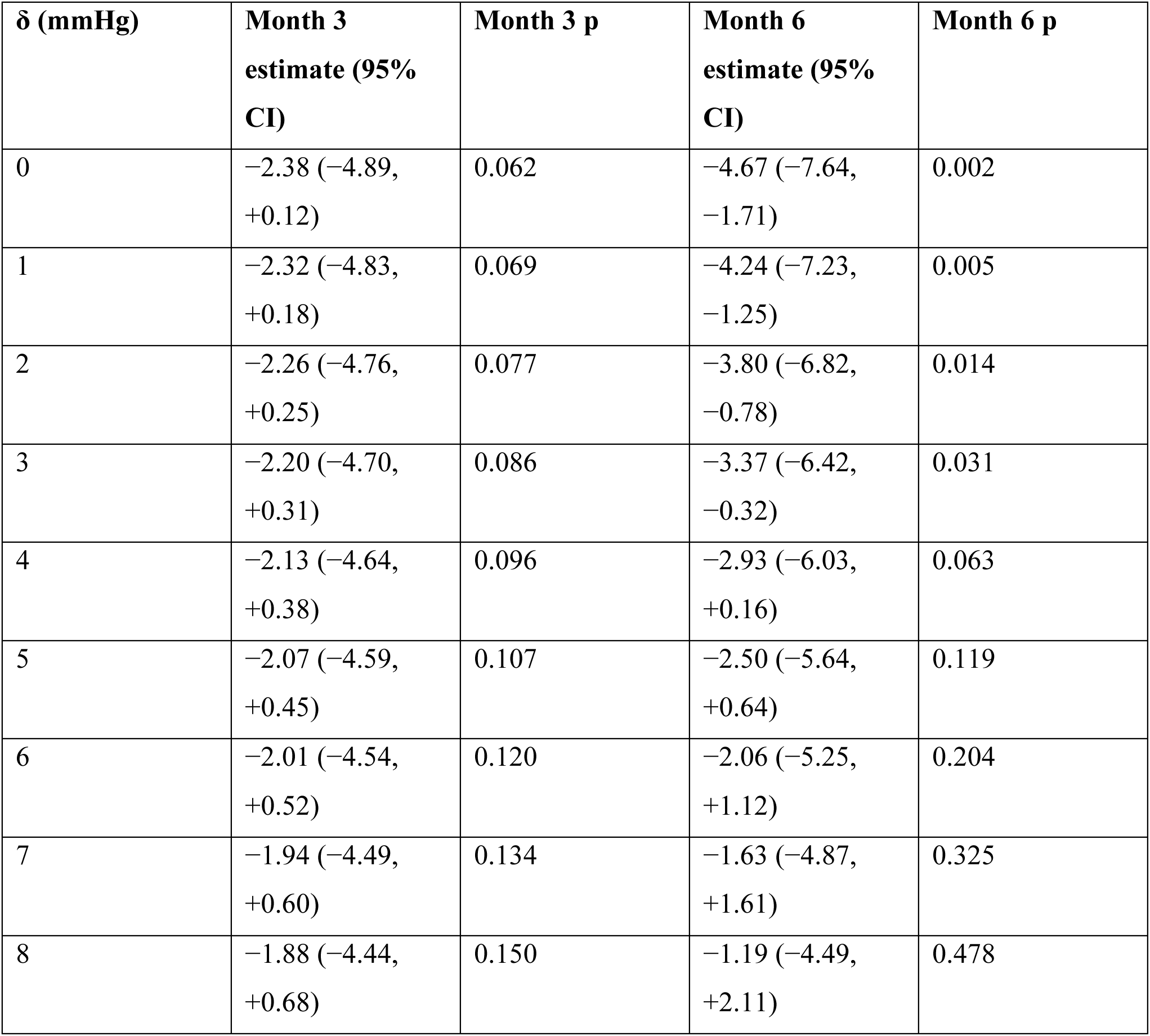
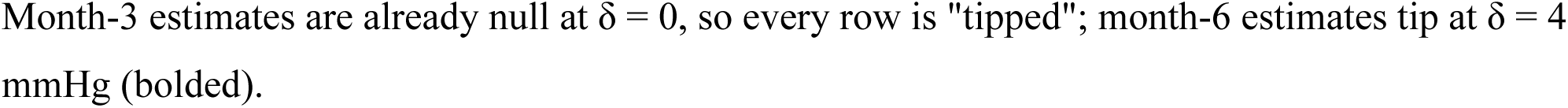
Values are the pattern-mixture model-based mean change in systolic blood pressure from baseline (mmHg, 95% CI) and its raw, unadjusted p-value, re-estimated at each erosion delta δ (0–8 mmHg) swept against every imputed (non-observed) follow-up value; δ = 0 is the base case, assuming missing-at-random. Larger δ represents a progressively larger assumed departure from missing-at-random, applied only in the direction that erodes the observed decline toward the null. The bolded row marks the tipping δ (the smallest value in the grid at which the 95% CI first includes zero), reported as a single figure in Table 3; this table shows the full erosion curve underlying it. CI, confidence interval; δ, erosion delta.

**SUPPLEMENTARY TABLE S8.**
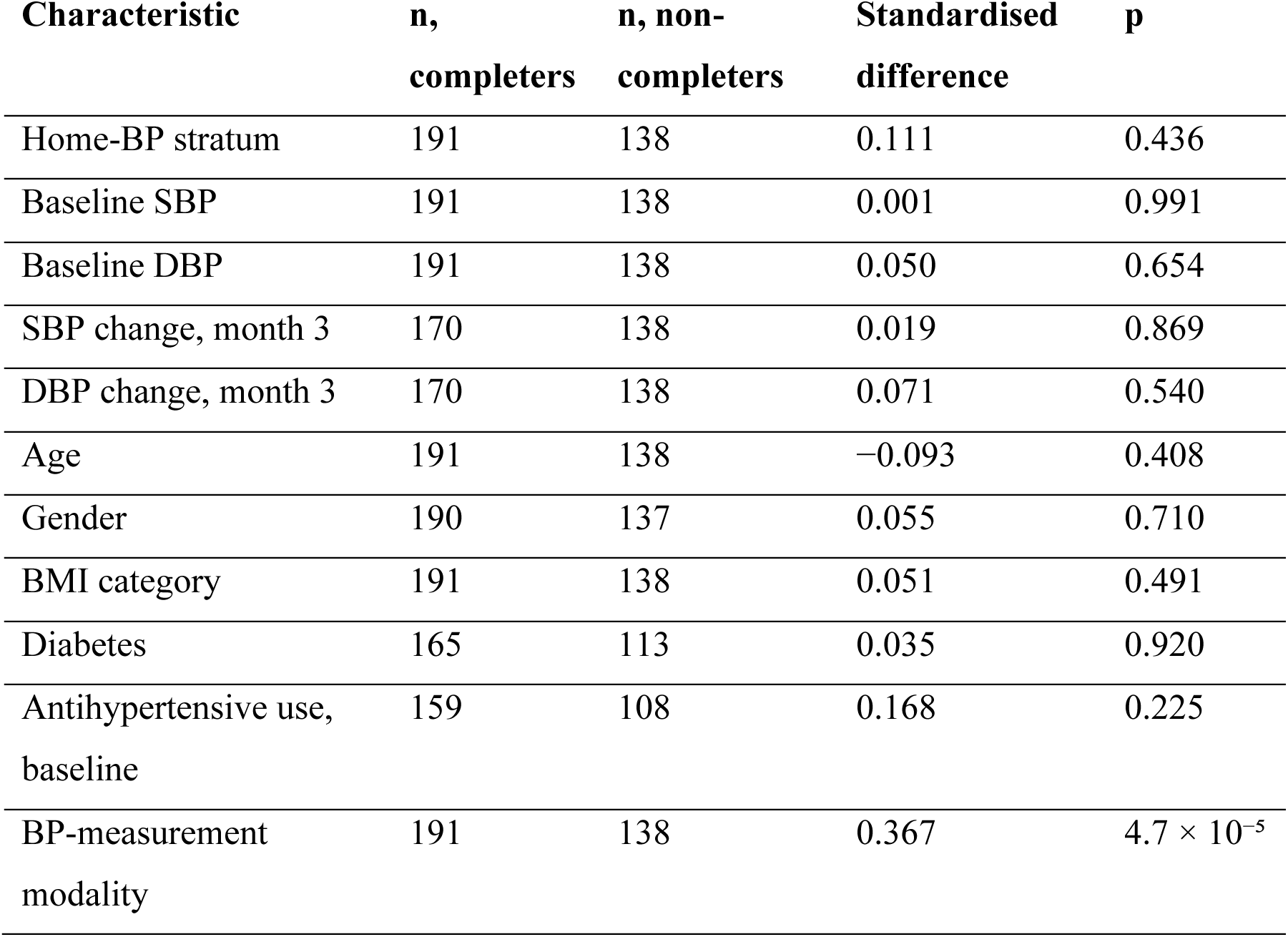
Attrition evidence for the: completers-versus-non-completers comparability. Compares the 191 members with sufficient readings in the month-6 window (149 6-month completers plus 42 whose window was still open at the data cutoff) against the 138 without, on baseline and early-outcome characteristics; standardised difference is the absolute mean difference in pooled SD units (categorical variables use Cohen’s h).

## 18 Supplementary Methods

### 18.1 AI-assisted meal rating and the nutrient-density flag-rate metrics

Dietary intake in this cohort is captured continuously through the platform’s in-app food-logging workflow, in which members photograph (or, optionally, describe in text) each meal. Photographs unsuitable for automated analysis are routed to a human nutrition coach; suitable photographs are passed to a proprietary AI image-analysis pipeline that classifies the foods present and estimates portion size. The platform combines this classification with internal per-food penalty rules covering inflammatory potential, fiber content and variety, and expected insulin response, and records, for each logged meal, whether it incurred an infraction on each of three categories: FIBER, INFLAMMATION, and INSULIN. The upstream image-recognition and rule-authoring methodology is proprietary and is described in greater detail in related work (33–38); this section describes only the elements needed to interpret the flag-rate results reported here.

The results reported in this manuscript use meal-level infraction flags directly, not the platform’s own point-weighted ND-score aggregate (nd_score_pct) or its per-axis severity values, neither of which is used anywhere in this manuscript. For each member and landmark window (the baseline and 3-/6– month windows defined in Methods), we computed four flag rates as the proportion of that member’s logged meals in the window meeting each definition: the share flagged on the FIBER axis, on the INFLAMMATION axis, on the INSULIN axis, and the share carrying no infraction on any axis (“clean” meals). These are frequency measures of how often a meal trips a category, not of how severe any single infraction is judged to be by the platform.

As with any platform-derived engagement metric, meal logging is opt-in and self-selected for motivation and engagement, and these flag rates are a coaching-prioritization proxy rather than a validated dietary-quality instrument: they are not benchmarked against established measures of dietary pattern such as the Healthy Eating Index, the Mediterranean Diet Adherence Screener, or the Alternative Healthy Eating Index, and they do not quantify nutrient magnitude or distinguish infraction severity, nor do they cover dietary domains (for example, sodium, alcohol, or whole-grain intake) that those instruments address.

