## Supplementary material for "Blood Pressure Reduction in Multiomic Digital Food-as-Medicine: a single-arm cohort study with a regression-to-the-mean-adjusted estimand": RECORD checklist

### RECORD Statement — Checklist of items, extended from STROBE, for studies using routinely collected health data

| # | Item | Where reported in the manuscript | Status |
|---|---|---|---|
| 1.1 | Type of data used should be specified in the title or abstract; name the databases if possible | Abstract, Methods sentence: "device-transmitted and self-reported home blood-pressure data" within a commercial digital food-as-medicine program; Methods names the specific device (Tenovi cellular monitor) and the phone-integration/manual-entry channels | Reported |
| 1.2 | Geographic region and timeframe in the title or abstract | Methods → Participants and digital health intervention: United States; enrolment window October 2025 – 31 August 2026 | Reported |
| 1.3 | If databases were linked, state so in the title or abstract | Methods: every variable derives from the program's own application/device data and a self-administered baseline questionnaire; no external database linkage is used in this analysis | Reported — stated as not applicable |
| 6.1 | Methods of study population selection, including codes/algorithms used to identify subjects | Data collection and analysis: index-date (t0) definition, baseline sufficiency rule, and the four hierarchically ordered analytic exclusion rules, given in full | Reported |
| 6.2 | Validation studies of the codes/algorithms used to select the population, if any | Data collection and analysis: the plausibility filter and aggregation approach are referenced to established home blood-pressure-monitoring guidance (Muntner et al., 2019; Stergiou et al., 2021) | Reported |
| 6.3 | If databases were linked, a flow diagram showing the linkage stages | Not applicable — no database linkage performed (see 1.3); Figure 1 covers the (non-linkage) selection flow | N/A |
| 7.1 | Complete list of codes/algorithms used to classify exposures, outcomes, confounders and effect modifiers | Data collection and analysis: blood-pressure outcome fully specified (filter bounds, occasion clustering, three-stage aggregation, window definitions, stratum cut-points); covariates defined via the self-administered questionnaire (Participants and digital health intervention) | Reported |
| 12.1 | Extent of investigators' access to the database population used to create the study population | Author Contributions and Conflict of Interest: investigators are employees of the program operator with direct roles (Data curation, Software, Methodology) over the underlying platform data used to construct the analytic cohort | Reported |

| # | Item | Where reported in the manuscript | Status |
|---|---|---|---|
| 12.2 | Data cleaning methods used | Data collection and analysis: physiological plausibility filter (with rationale against distribution-based trimming), first-calendar-day retention decision (with its own empirical check reported in Results), and occasion-clustering rule | Reported |
| 12.3 | Whether the study involved person-level, institutional-level, or other data linkage across two or more databases; linkage methods and quality evaluation | Not applicable — no cross-database linkage, consistent with 1.3 and 6.3 | N/A |
| 13.1 | Detailed description of the selection of persons included in the study (filtering on data quality, data availability, linkage), by text and/or flow diagram | Figure 1 and Results, opening paragraph: eligibility funnel from assessed population to analysed cohort, with per-reason exclusion counts | Reported |
| 19.1 | Discuss the implications of using data not created to answer the specific research question — misclassification, unmeasured confounding, missing data, changing eligibility over time | Discussion: misclassification (screening vs diagnostic staging, κ and specificity figures cited); unmeasured confounding (intake-only medication capture); missing data (attrition limitation and the three-estimator sensitivity approach) | Reported |
| 22.1 | How to access supplemental information: study protocol, raw data, or programming code | Ethics Statement (IRB protocol E&I Review Services #18053); Data Availability Statement (member-level data withheld under consent restrictions, available on reasonable credentialed request); Code Availability Statement (custom analysis code available from the corresponding author upon reasonable request). | Reported |
