## Supplementary material for "Blood Pressure Reduction in Multiomic Digital Food-as-Medicine: a single-arm cohort study with a regression-to-the-mean-adjusted estimand": STROBE checklist

### STROBE Statement — Checklist of items to include when reporting observational studies (cohort studies)

| # | Item | Where reported in the manuscript | Status |
|---|---|---|---|
| 1a | Study design indicated in title or abstract with a commonly used term | Title ("…a single-arm cohort study…"); Abstract, Methods sentence ("retrospective, single-arm cohort study…") | Reported |
| 1b | Informative and balanced abstract summarising what was done and found | Abstract (Background, Methods, Results, Conclusion): design, N, strata, follow-up, unadjusted and RTM-corrected estimates with 95% CI and p for both target strata | Reported |
| 2 | Background/rationale for the investigation | Introduction ¶¶1–3: cardiovascular risk gradient of SBP reduction, HBPM guideline status, prior single-arm literature and its uncorrected estimates, RTM named as the validity threat, cost-effectiveness context | Reported |
| 3 | Objectives, including any prespecified hypotheses | Methods → Statistical analysis → Models and estimands: primary estimand (unadjusted paired change, 6 months, Stage 1/Stage 2) and secondary estimand (RTM-corrected ANCOVA) explicitly named; at-goal stratum "designated in advance of estimation" as the negative control | Reported |
| 4 | Key elements of study design presented early | Methodology → Study design and setting: design and primary/secondary outcomes stated up front | Reported |
| 5 | Setting, locations, and relevant dates (recruitment, exposure, follow-up, data collection) | Setting (US, commercial digital program); enrolment window (October 2025 – 31 August 2026); landmark windows (3, 6 months, ±30 days) | Reported |
| 6a | Eligibility criteria and sources/methods of selection; follow-up methods | Participants and digital health intervention (program-level eligibility); Data collection and analysis (index date, hierarchical exclusion order, four exclusion reasons, baseline/landmark window definitions) | Reported |
| 6b | (Matched studies) matching criteria and numbers exposed/unexposed | Discussion: at-goal stratum used as internal negative control rather than a matched comparator; explicitly discussed as such | N/A — single-arm design, no matched comparator |
| 7 | Definitions of outcomes, exposures, predictors, confounders, effect modifiers | Methods: outcomes (home SBP/DBP change), strata (at-goal/Stage 1/Stage 2), covariates (age, BMI category, T2D, CKD, antihypertensive use), | Reported |

| # | Item | Where reported in the manuscript | Status |
|---|---|---|---|
| | | effect modifier (measurement modality) | |
| 8 | Data sources and measurement methods for each variable | Data collection and analysis: Tenovi cellular monitor; phone-integration and manual-entry channels; plausibility filter (SBP 70–260 / DBP 40–160); three-stage occasion→day→window aggregation | Reported |
| 9 | Efforts to address potential sources of bias | Introduction (RTM named explicitly); Methods (at-goal internal negative control, ANCOVA correction); Sensitivity analyses (season, modality, drop-one covariates, alternative RTM slope) | Reported |
| 10 | Study size, and how it was arrived at | Figure 1 (full eligibility funnel, eligible population → 329 analysed); Results (realised n per stratum × landmark cell) | Reported |
| 11 | Handling of quantitative variables; groupings and why | Methods (home-BP strata with numeric cut-points and office equivalents); Sensitivity analyses (WHO BMI categories with cut-points) | Reported |
| 12a | Statistical methods, including confounding control | Statistical analysis → Models and estimands: reduced ANCOVA-LMM fully specified (equation, every term defined), REML, R/lme4/emmeans named | Reported |
| 12b | Subgroups and interactions | Sensitivity analyses: antihypertensive-use and T2D subgroups with CIs; stratum × baseline-slope interaction with likelihood-ratio test | Reported |
| 12c | Missing data | Sensitivity analyses: pattern-mixture/MICE, IPCW, and naive complete-case estimators reported side by side | Reported |
| 12d | (Cohort) how loss to follow-up was addressed | Sensitivity analyses: same three-estimator approach; tipping-point analysis; completer-vs-non-completer comparison (Supplementary Table S8) | Reported |
| 12e | Sensitivity analyses | Sensitivity analyses: modality, covariate drop-one, season, stratum-slope interaction, alternative RTM anchor, attrition | Reported |
| 13a | Numbers of individuals at each stage (eligible, examined, confirmed eligible, included, analysed) | Figure 1 (participant flow); Results, opening paragraph | Reported |
| 13b | Reasons for non-participation at each stage | Figure 1 caption; Results, opening paragraph — each exclusion category carries a count | Reported |

| # | Item | Where reported in the manuscript | Status |
|---|---|---|---|
| 13c | Consider use of a flow diagram | Figure 1 | Reported |
| 14a | Participant characteristics and information on exposures/confounders | Table 1: age, sex, baseline BP, BMI, T2D, CKD, antihypertensive use, biomarkers, BP-source modality, by stratum and overall | Reported |
| 14b | Number of participants with missing data for each variable | Table 1 (per-marker biomarker denominators, per-cell n); Supplementary Table S8 (completers vs non-completers) | Reported |
| 14c | (Cohort) summarise follow-up time | Supplementary Table S1: monitoring intensity and median day of last reading per stratum × landmark cell | Reported |
| 15 | (Cohort) outcome events or summary measures over time | Table 2: unadjusted and RTM-corrected estimates with 95% CI, p and n for every stratum × landmark × pressure-type cell | Reported |
| 16a | Unadjusted and (if applicable) adjusted estimates with precision | Table 2 pairs every unadjusted estimate with its RTM-corrected counterpart; Sensitivity analyses adds the fully-adjusted-covariate estimates | Reported |
| 16b | Category boundaries when continuous variables were categorised | Methods (home-BP stratum cut-points); Sensitivity analyses (WHO BMI category boundaries) | Reported |
| 16c | Translate relative risk into absolute risk where relevant | Discussion: corrected 6-month Stage-2 decline anchored to the Ettehad (2016) and Blood Pressure Lowering Treatment Trialists' Collaboration (2021) cardiovascular-risk meta-analyses | Reported |
| 17 | Other analyses (subgroups, interactions, sensitivity) | Sensitivity analyses (modality, covariates, season, alternative RTM anchor, attrition); Weight and blood pressure change (Table 4, Mundlak within/between-person decomposition); meal-level dietary flag-rate metrics and blood pressure change (Table 5, same decomposition); Internal negative control subsection (reliability-based RTM cross-check) | Reported |
| 18 | Key results, with reference to objectives | Discussion, opening paragraph: restates the unadjusted and RTM-corrected estimates against the study objective | Reported |
| 19 | Limitations: sources of bias/imprecision, direction and magnitude | Discussion, limitations paragraph: single-arm/no concurrent control; RTM magnitude; screening vs diagnostic staging; self-selection/generalisability; intake-only medication capture; the negative- | Reported |

| # | Item | Where reported in the manuscript | Status |
|---|---|---|---|
| | | control stratum's own medicated status; unmeasured mediators; no external comparator | |
| 20 | Cautious overall interpretation | Association language used throughout ("was associated with"); Conclusion: "no causal inference is warranted from this design" | Reported |
| 21 | Generalisability of the study results | Discussion and limitations: self-selected engaged home monitors, commercial program, employer-sponsored benefits, obesity-based eligibility, screening classification | Reported |
| 22 | Funding source and role of funders | Funding: Digbi Health named as funder; Conflict of Interest: employment, equity, CEO/founder role, royalties, pending patents disclosed by named author | Reported |
